# Informed Injury Prediction in Elite Football: Decision Theory meets Machine Learning

**DOI:** 10.1101/2025.04.23.25326218

**Authors:** Manuel Huth, Berta Canal-Simón, Eva Ferrer, Gil Rodas, Xavier Yanguas, Jan Hasenauer, Juan R González

**Affiliations:** Life and Medical Sciences (LIMES) Institute, University of Bonn, Bonn, Germany; Bonn Center for Mathematical Life Sciences, University of Bonn, Bonn, Germany; Barcelona Institute for Global Health (ISGlobal), Barcelona, Spain; Made of Genes, Barcelona, Spain; Medical Department of Football Club Barcelona (FIFA Medical Centre of Excellence), and Barça Innovation Hub of Football Club Barcelona, Barcelona, Spain; Sports and Exercise Medicine Unit, Hospital Clinic and Sant Joan de Déu, Barcelona, Spain; Leitat technological Center, Terrassa, Spain; CIBER in Epidemiology and Public Health (CIBERESP), Barcelona, Spain; Department of Mathematics, Universitat Autònoma de Barcelona (UAB), Barcelona, Spain

## Abstract

Injuries in elite sports disrupt team performance, shorten careers, and incur significant financial costs, highlighting the critical need for accurate predictions to inform optimal decisions that effectively prevent injuries. Existing approaches to injury prediction fail to account for cumulative risk, overlook injury severity, lack reliable probability calibration, and omit statistically guided decision thresholds. Here, we present a novel injury prediction framework integrating risk accumulation via survival analysis with machine learning, probability beta calibration, and statistical decision theory. Using a unique dataset spanning four seasons from FC Barcelona’s women’s team, we demonstrate that our framework outperforms standard classifiers, yielding superior discrimination ability. Our framework identifies fatigue-related measures as key injury predictors and incorporates flexible thresholds based on match importance and decision-maker certainty, improving player availability. Scalable and transferable to other sports, this framework bridges academic research and practical deployment, empowering sports organizations to optimize player performance and long-term outcomes.

## Introduction

Injuries in elite sports pose a significant challenge with wide-ranging implications, including disruptions to club performance, shortened player careers, and destabilized long-term financial sustainability^1–4^. During the 2023/24 season alone, 4,123 injuries were recorded across the top five European leagues (England, Spain, Italy, Germany, and France), resulting in a financial burden of around 732.02 million euros^5^.

Efforts to predict daily injury risk using player tracking data and machine learning have emerged as a key strategy for mitigating these impacts^6–9^. These approaches leverage predictive features, such as training loads, sleep quality, demographics, or psychological factors to anticipate injuries. Yet, significant challenges remain: a recent review by Bullock et al.^10^ found that most existing models lack adequate performance assessment, probability calibration, and transparency, rendering them unsuitable for practical deployment. Similarly, Leckey et al.^11^ highlight the misalignment between methodological development and practical applicability, emphasizing that research teams must integrate both technical and domain expertise as even robust machine learning models are ineffective if they fail to address the real needs of practitioners.

We identify three additional gaps that hinder current injury prediction frameworks. First, existing machine learning classifier-based approaches^6–9;12;13^ overlook the cumulative nature of injury risk. Specifically, these methods fail to capture how injury probability increases monotonically with the duration of exposure during training or matches. For example, a player participating for 70 minutes should inherently face a lower injury risk than one playing for 90 minutes. Second, models lack day-specific tailored decision thresholds that reflect the varying importance of matches versus training sessions. For example, a player with moderate injury risk might strategically rest during a training day but participate in a match due to its higher strategic and financial value. Constructing these thresholds requires reliable probability calibration—that is, ensuring that predicted probabilities align with observed outcomes. For example, if a model predicts a 5% chance of injury, then approximately 5% of those cases should indeed result in an injury^14^. Unfortunately, traditional machine learning models often fail to achieve this level of calibration without further adjustments ^15^. Consequently, any robust framework must include mechanisms to correct miscalibrated probabilities. Third, commonly used metrics like Recall^6–8^ or the F1 score^6;9^ fail to account for the severity of injuries, measured in injury duration. While detecting a single long-term injury may be highly beneficial for team performance and availability, traditional evaluation metrics penalize false positives without incorporating the practical impact of injury severity.

To address these limitations, we introduce a novel injury prediction framework that integrates survival analysis with machine learning, probability calibration, and practical decision-making strategies derived from statistical decision theory - all designed to meet the real-world needs of practitioners. Specifically, our approach models injury risk by treating each activity as a survival observation, enabling the estimation of injury probabilities over time using methods from survival analysis that naturally account for injury risk accumulation over time^16–18^. We combine GPS-derived training load features to create a comprehensive and individualized assessment of injury risk. To ensure that predicted probabilities are reliable for decision-making, we apply beta calibration^19^, a technique widely used in fields such as oncology research where accurate probabilities are essential^20;21^. Furthermore, we derive actionable decisions by incorporating day-specific valuations for matches and training sessions, alongside a pre-specified level of rest benefit certainty (RBC) that reflects decision-makers’ required confidence to rest a player.

We evaluate our framework using a dataset spanning four seasons (2019/20 to 2022/23) from Fútbol Club Barcelona’s (FC Barcelona, Spain) first women’s team^22;23^. Injuries are considered to reflect a team’s time loss of player availability. Injury evaluations were conducted by the team’s medical physician in collaboration with the FC Barcelona medical department, adhering to standardized diagnosis and return-to-play protocols established in the club’s guidelines^24;25^. The analysis specifically targeted non-contact injuries involving muscles, tendons, ligaments, and cartilage. First, we assess the robustness of survival-based models compared to classical machine learning classifiers ^26–28^ over 400 training, calibration, and test splits from the 2019/20, 2020/21, and 2021/22 seasons. Model performance is evaluated using a player availability gain metric, which accounts for injury durations to provide a practical assessment of prediction impact. Second, we identify the most important predictive features across these splits to better understand key contributors to injury risk. Finally, we validate the best-performing model and selected features on the previously unseen 2022/23 season using a rolling prediction approach, simulating real-world conditions for decision-making.

## Results

### Integrating Risk Accumulation, Calibration, Match Valuations and Decision Certainty in Injury Prediction

An effective injury prediction framework for clubs, particularly for coaches and medical staff to which we refer as team decision-makers, must meet three criteria: (1) capture the non-linear accumulation of injury risk over training time, (2) provide accurate probability scores reflecting true injury risk, and (3) offer interpretable thresholds to translate probabilities into actionable decisions, such as predicting to rest a player or to let her play.

In this study, we introduce an injury prediction framework that meets these criteria by incorporating risk accumulation over time and dynamically adjusting decision thresholds based on match and practice session importance as well as required certainties of decisions: (1) We interpret the machine learning task as a daily survival analysis for each player (Figure 1A). Survival in this context refers to the time until an injury in minutes. Each activity day represents a survival observation, with played minutes treated as the time indicator and the injury label acting as the censoring indicator. Unlike traditional methods that treat this as a pure classification task^6;13;29^, survival analysis captures the dynamics of injury risk accumulation over the activity time by a monotonic decreasing survival curve. As main survival model, we use the Extreme Gradient Boosting Survival Embedding (XGBSE) ^16^. For further comparison, we also include the Accelerated Failure Time ^17^, Coxnet^18^, and boosted Cox^30^ survival models. The machine learning survival model is trained using the input features, e.g. accumulated GPS data, a match indicator and age, on the training set. This model generates survival curves for unseen observations based on these input features (Figure 1B). Survival curves evaluated at the planned training duration yield an estimate of the probability of sustaining no injury up to the planned training duration, which is then used to compute the probability of injury given the planned training duration. By construction of the monotone survival curves, longer training durations naturally lead to higher injury risk, reflecting risk accumulation over time. In contrast, standard classification models like standard Light- GBM^26^, which treat played minutes as an ordinary feature, fail to reflect this cumulative risk. For LightGBM, a monotonization feature has been introduced that could, in principle, deal with the monotone risk accumulation. We benchmark the monotone feature and other standard machine learning classifier^26–28^ against the survival analysis approach within our results. For model training, calibration and evaluation, the data is split into a training, a calibration, and a test set.

**Figure 1:**
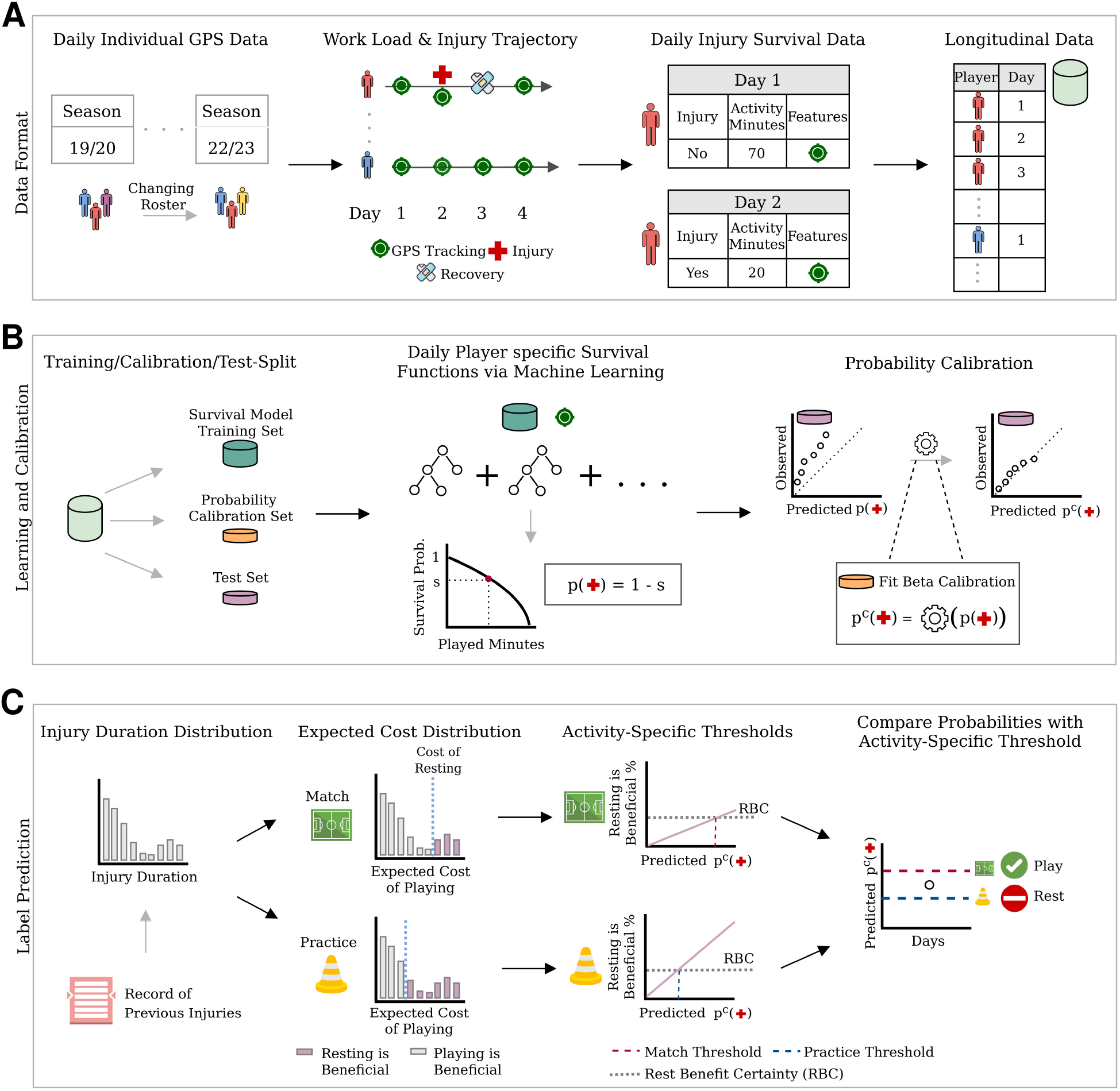
Data Format, Learning and Label Prediction. (**A**) Data Format: Daily GPS-based training load and injury records were collected over 4 seasons with a changing player roster. Each day is treated as an individual survival observation, where the injury status serves as the censoring indicator and activity minutes as survival time. Combining daily observations per player creates a longitudinal survival dataset. (**B**) Learning and Calibration: The data is split into training, calibration, and test sets. The training set is used to fit the survival model, which outputs the probability of injury given planned activity time. The calibration set is used to refine injury probabilities via beta calibration using observed injury indicators. (**C**) Label Prediction: Injury distributions are estimated from past records. For match and practice days, expected costs of resting and playing are compared for each historical injury duration. If the Rest Benefit Certainty (RBC), e.g., 50%, favors resting, the player is predicted to rest; otherwise, they play.

(2) To align predicted probability scores with the actual injury risk, we calibrate predicted probabilities using a separate calibration set and a beta calibration model ^19^. This calibration ensures that the reported probabilities reflect actual injury risk and thus are actionable for team decision-makers (Figure 1B). Without calibration, thresholds for actionable decisions may become unreliable, leading to overly conservative or risky choices. For instance, miscalibrated probabilities might underestimate injury risk, exposing players to harm, or overestimate it, sidelining key players unnecessarily. Beta calibration is particularly suitable due to its robustness to skewed probability distributions^19^. We validate the calibration for our data by comparing the calibrated injury probabilities against the uncalibrated probabilities.

(3) Calibrated probabilities provide a robust foundation for playing-or-resting decision-making but must be paired with actionable decision thresholds. Our framework leverages pre-specified activity valuations that can be tailored to individual players and specific days. These valuations reflect the importance of participation on any given day, enabling decisions that align with team priorities. A higher valuation is reflected in a higher probability threshold required to rest a player. For instance, these valuations can reflect the higher stakes of critical matches or lower priority of routine training (Figure 1C).

Furthermore, coaches and medical staff can specify a minimum certainty level required to rest a player - reflecting their risk preferences. This is achieved by calculating the expected costs of playing - which depend on the distribution of potential injury durations - and comparing them to the fixed cost of resting. This comparison quantifies the likelihood that resting a player will result in a player Availability Gain (AG). For example, based on predicted injury risk and observed injury durations, decision-makers might agree on resting a player if it is beneficial in 50% of injury duration scenarios. Decision-makers can use this level of certainty to guide their actions, which we label as Rest Benefit Certainty (RBC).

To introduce these match valuation and RBC concepts further to the reader, we have included examples with concrete numbers in the Supplementary Material.

### Collection, Assembly and Curation of Training Data

Our study compiled a dataset for FC Barcelona’s first women’s team over four consecutive seasons (2019/2020 to 2022/2023). This integrated dataset combines daily training load records, match schedule information, injury reports, and international duty data. When training data were missing due to players’ international duty, match participation (e.g., minutes played) was used for imputation of training load. Daily injury records - detailing occurrence and duration - further enriched our dataset, allowing us to examine links between training, performance, and injury risk. All data were harmonized to a common timeline, providing a robust foundation for our analysis.

The dataset includes 34 players, with turnover as some players left and new players joined the club during this period (Figure 2A). This turnover introduces variability due to unobserved player differences, which could affect model performance and requires results to be interpreted accordingly. The players’ ages ranged from 17 to 33 years, with a slight year-to-year increase due to natural aging (Figure 2B). This increase in ages was offset by younger players joining the team in 2022/23, resulting in marginal changes to the age distribution. The frequency of matches and practices varied over time due to injuries, international duties, personal breaks, and the COVID-19 disruption during the 2019/20 season. While practices outnumbered matches (Figure 2C), the distribution of covered kilometers per day was stable across seasons and higher for matches compared to practices (Figure 2D). These factors were incorporated into the survival model using a match indicator and aggregated training load variables from days prior to the day of activity. Over four seasons, 83 non-contact injuries were recorded (0.62% of all data points), with a pronounced spike in 2021/2022 (31 injuries) compared to the prior season (14 injuries; Figure 2E). It is striking that injuries occur at a high rate during matches, even though the total practice time per season is approximately four times greater than the match time (Supplementary Figure S1) suggesting an increased non-contact injury risk during matches. The high class imbalance and the shift in injury label distribution present significant challenges for injury prediction models as these fluctuations can undermine the stability and predictive performance of models trained on historical data. Injury durations ranged widely, with a mean of 37.8 days and a median of 23 days (Figure 2F). This disparity highlights the influence of long-term injuries, such as ligament injuries, and importance of analyzing the full injury duration distribution, rather than relying on averages, for informed decision-making.

**Figure 2:**
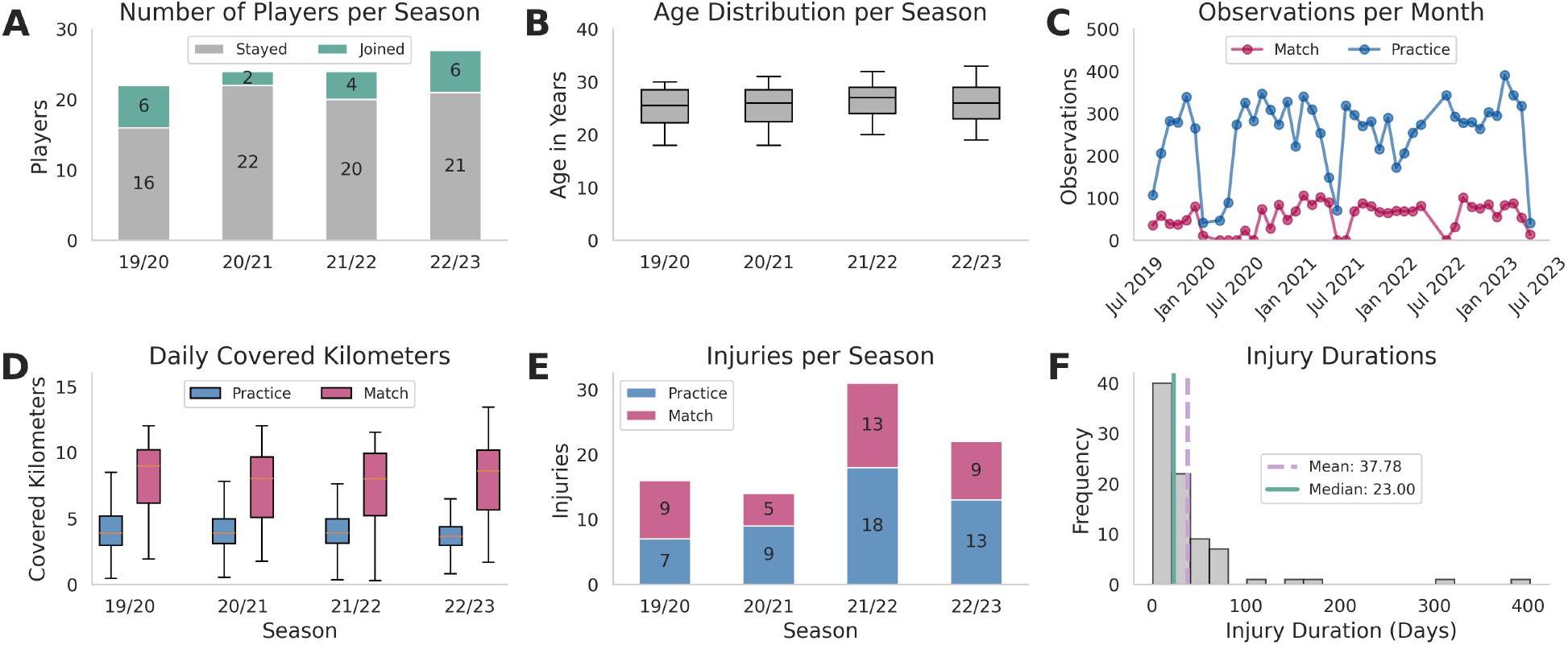
Summary of player characteristics, observations, and injuries across seasons. **(A)** Number of players per season, with returning players shown in gray and new players in green. **(B)** Boxplot of age distribution of players per season. **(C)** Monthly number of observations, split into matches (red) and practices (blue). **(D)** Boxplot of daily covered kilometers, shown as boxplots. **(E)** Number of injuries per season, separated into match injuries (red) and practice injuries (blue). **(F)** Distribution of missed days per injury.

### Survival Models Outperform Standard ML Classifiers in Discrimination Ability

To evaluate the discrimination ability of our proposed framework, we compared its performance against alternative survival models and standard machine learning classifiers. Here, discrimination refers to a model’s capacity to distinguish between positive (injury) and negative (no injury) cases independently of any decision threshold. This approach enables us to assess model performance without the confounding effects of threshold selection. For comparison, we have chosen LightGBM^26^ as tree-based Extreme Gradient Boosting has performed best in previous injury prediction attempts^7–9^, and additionally k-nearest-Neighbors^27^ and Logistic Regression^28^ for a broader scope of the comparison. We also incorporate two additional variants of LightGBM. The first enforces a monotonic relationship for playing time so that higher training loads correspond to increased injury risk. The second leverages Light-GBM as a tree embedding, which is then fed into a Logistic Regression model—mimicking the approach behind XGBSE for a standard machine learning classifier.

We quantified discrimination using the Mean Average Precision (MAP) and the Area Under the Receiver Operating Characteristic Curve (AUROC). While AUROC has been widely employed in injury prediction studies^6–8^, Mean Average Precision (MAP)—which jointly accounts for both precision and recall—has received relatively little attention. This is despite its proven effectiveness in handling highly imbalanced datasets^31;32^, a common challenge in injury prediction where healthy player records far exceed those of injuries. As a baseline, we employed a prior mean classifier that leverages the inherent class frequencies to ensure meaningful comparisons. Data were partitioned using a month-wise block splitting strategy (Figure 3A) to preserve temporal dependencies. This approach split the data into distinct training, calibration, and test sets, and the splitting process was repeated 400 times to ensure robust performance estimates.

**Figure 3:**
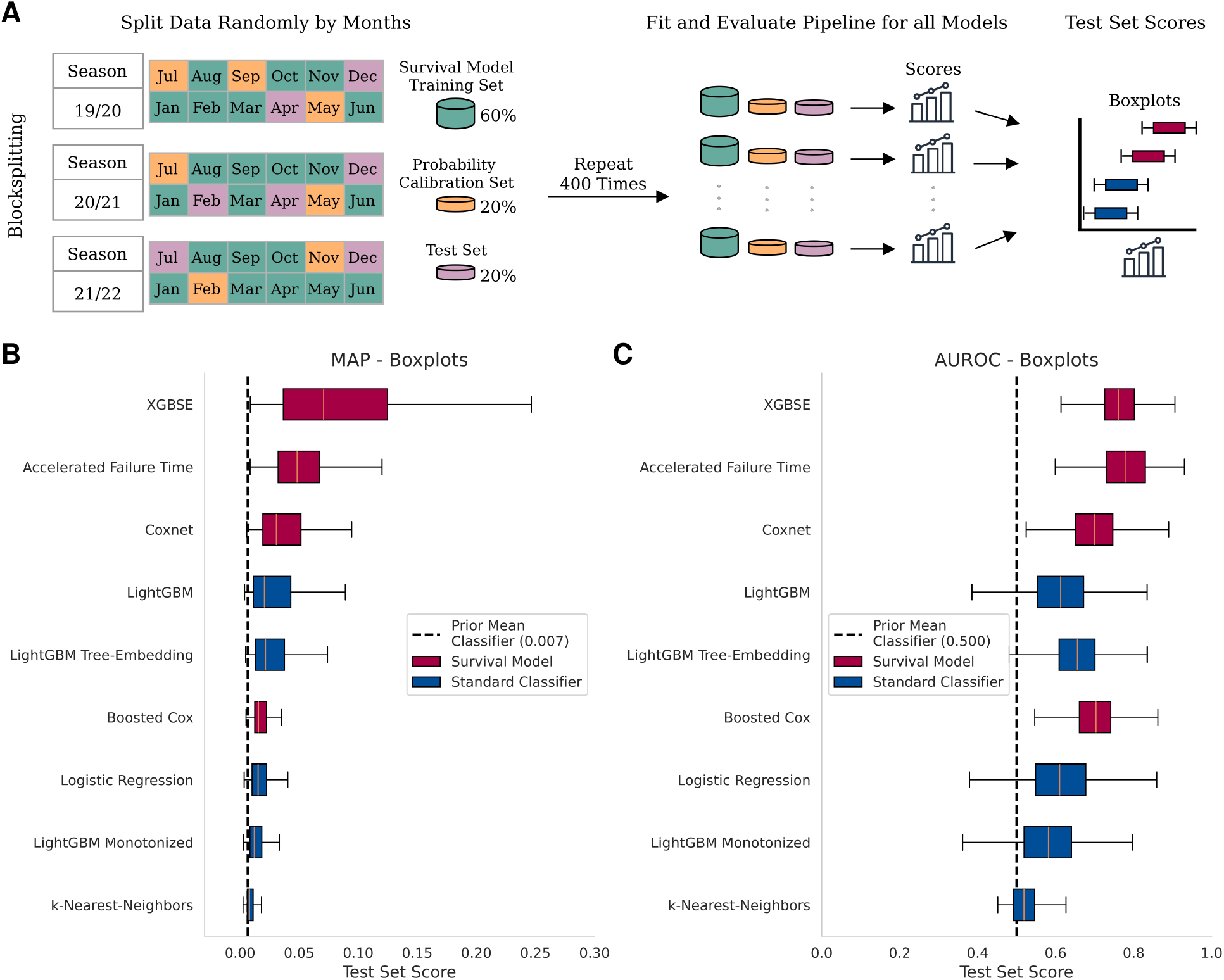
Discrimination Performance. Survival-based models are shown in red, and standard machine learning classifiers in blue. **(A)** The sample is split into training, calibration, and test sets, repeated 400 times with different seeds using a month-wise block-splitting such that observations from the same month stay within the same set. For each split, the framework is fitted, and the metrics are computed. **(B, C)** Boxplots of Mean Average Precision (MAP) and Area Under the Receiver Operating Characteristic Curve (AUROC) across 400 splits.

XGBSE achieved a median MAP of 0.071 - an approximately 11-fold improvement over the baseline mean classifier (Figure 3B) - and a median AUROC of 0.761, improving the baseline mean classifier substantially.

In further comparisons, XGBSE outperformed the best standard classifier, LightGBM (which achieved a median MAP of 0.021), by a factor of roughly 3.44. Although the Accelerated Failure Time model demonstrated the second-best MAP (median: 0.048) and the highest AUROC (median: 0.780), its performance was closely followed by XGBSE in terms of AU-ROC. The similarity in performance between these models is unsurprising given that XGBSE leverages the Accelerated Failure Time model as a feature transformer.

Overall, our results show for both considered metrics that survival models, particularly XGBSE, offer superior discrimination ability compared to standard machine learning classifiers.

### Beta Calibration Aligns Predicted Probabilities with True Injury Risk

To assess if the models provide interpretable probability estimates that are well-aligned with actual injury risk, we analyzed how the predicted probabilities generated by XGBSE aligned with the observed proportions of injuries. By performing calibration on an independent calibration set - distinct from both the training and test sets - our approach ensures an unbiased alignment of predicted probabilities with the true injury risk for each of the 400 splits (Figure 4A). We computed average calibration curves by deriving a calibration curve for each split and then taking their point-wise average.

**Figure 4:**
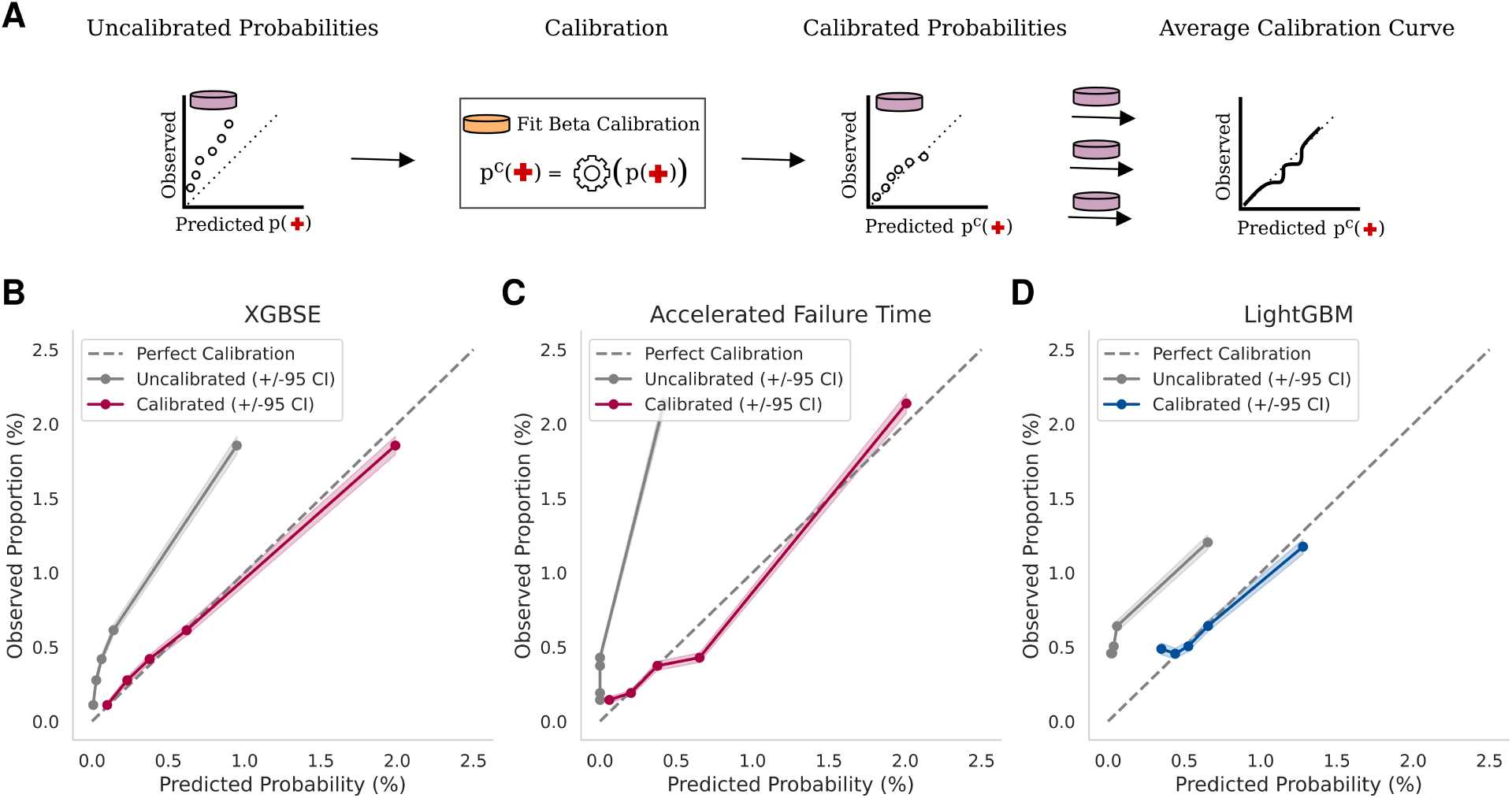
Calibration Performance. Average calibration curves for uncalibrated and calibrated predictions, using 5 quantile bins per curve. Averages are computed point-wise across 400 splits, with the 45° line indicating perfect calibration. **(A)** Representation of calibration workflow. **(B)** For the XGBSE model. **(C)** For the Accelerated Failure Time model. **(D)** For LightGBM.

Our analysis showed that the uncalibrated XGBSE model systematically underestimated the observed proportions of injuries, as evidenced by the average calibration curve (Figure 4B). After calibration, the average calibration curve closely aligned with the observed injury rates, demonstrating improved accuracy and reliability in the predicted probabilities.

We observed similar behavior with other methods, such as the Accelerated Failure Time survival model (Figure 4C) and LightGBM (Figure 4D), demonstrating that calibration generally improves risk alignment. However, these results, combined with our earlier findings on discrimination ability, indicate that while beta calibration can effectively align risk estimates, a robust underlying model is essential for achieving strong predictive power. For the survival models, calibration involved only monotone transformations that did not alter the MAP or AUROC while for LightGBM, only some transformations were non-monotone (Supplementary Figure S2) such that results stay qualitatively the same.

### Uncertainty-Aware Cut-offs Improve Player Availability Across Match Valuations

After examining the discrimination and calibration capabilities of our proposed framework, we evaluated its performance in decision-making scenarios to test its applicability as decision support for coaches and medical staff (Figure 5A). Specifically, we analyzed three match valuation scenarios: *R* = 2, 5, 10. Here, *R* represents the relative match valuation, quantifying how much more important a match is compared to a training session. The calculations were conducted for five RBC thresholds ranging from 50–90%, representing varying levels of confidence required to rest a player. The primary evaluation metric, Mean Availability Gain (MAG), quantifies the trade-off between the costs of unnecessary rest and the benefits of detecting injuries that lead to lost match and practice days. A more detailed explanation is provided in the Methods section.

**Figure 5:**
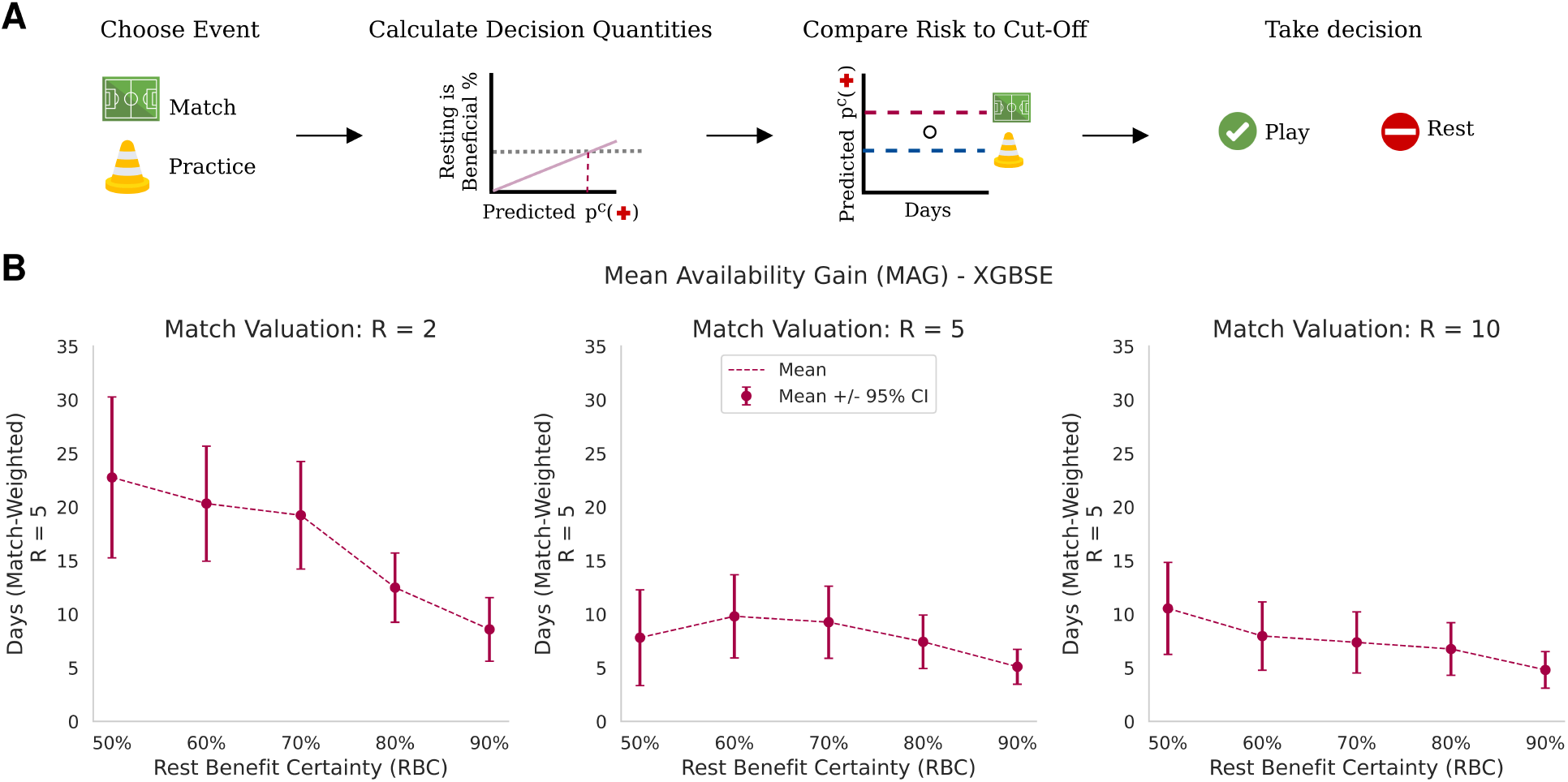
Rest Benefit Analysis of Player Availability. **(A)** Representation of Decision Process. **(B)** Mean Availability Gain as a function of Rest Benefit Certainty (RBC) and match valuation, calculated over 400 samples for the XGBSE model. Confidence intervals represent the 95% confidence range for the mean, obtained via a normal approximation.

Each of the split results were multiplied by a scaling factor to match the number of observations from the 2022/2023 season making the results comparable with the seasonal analysis of 2023. Across all match valuations and RBC thresholds, we observed consistently positive MAGs (Figure 5B).

Specifically, a RBC range of 50–70% consistently yielded the highest MAGs across all match valuations. For higher match valuations, the differences in MAGs across RBCs were smaller, suggesting greater flexibility in threshold selection. Notably, MAGs within the 50–70% RBC range were not statistically different from each other at the 95% confidence level across all match valuations.

We analyzed the precision, recall and the F1-score, to evaluate the trade-offs in predicting injuries within a highly imbalanced dataset while ignoring the impact of injury durations (Supplementary Figure S3). For a match valuation of 5 and a RBC of 60%, precision indicates that around 1 in 11 (9.1%) predicted rest days corresponded to actual injuries, while recall shows the model detected around 1 in 12 (8.1%) injury events.

### Cumulative Load Emerges as the Strongest Predictor

To better understand the mechanics of our method’s predictive performance and to assess feature relevance, we examined the influence of individual features on predictions (Figure 6). Specifically, we calculated two metrics for each sample split: the average gain in the loss function and the frequency with which each feature was selected for splits across all trees in the ensemble. The medians across all sample splits were used to summarize results (Figure 6A).

**Figure 6:**
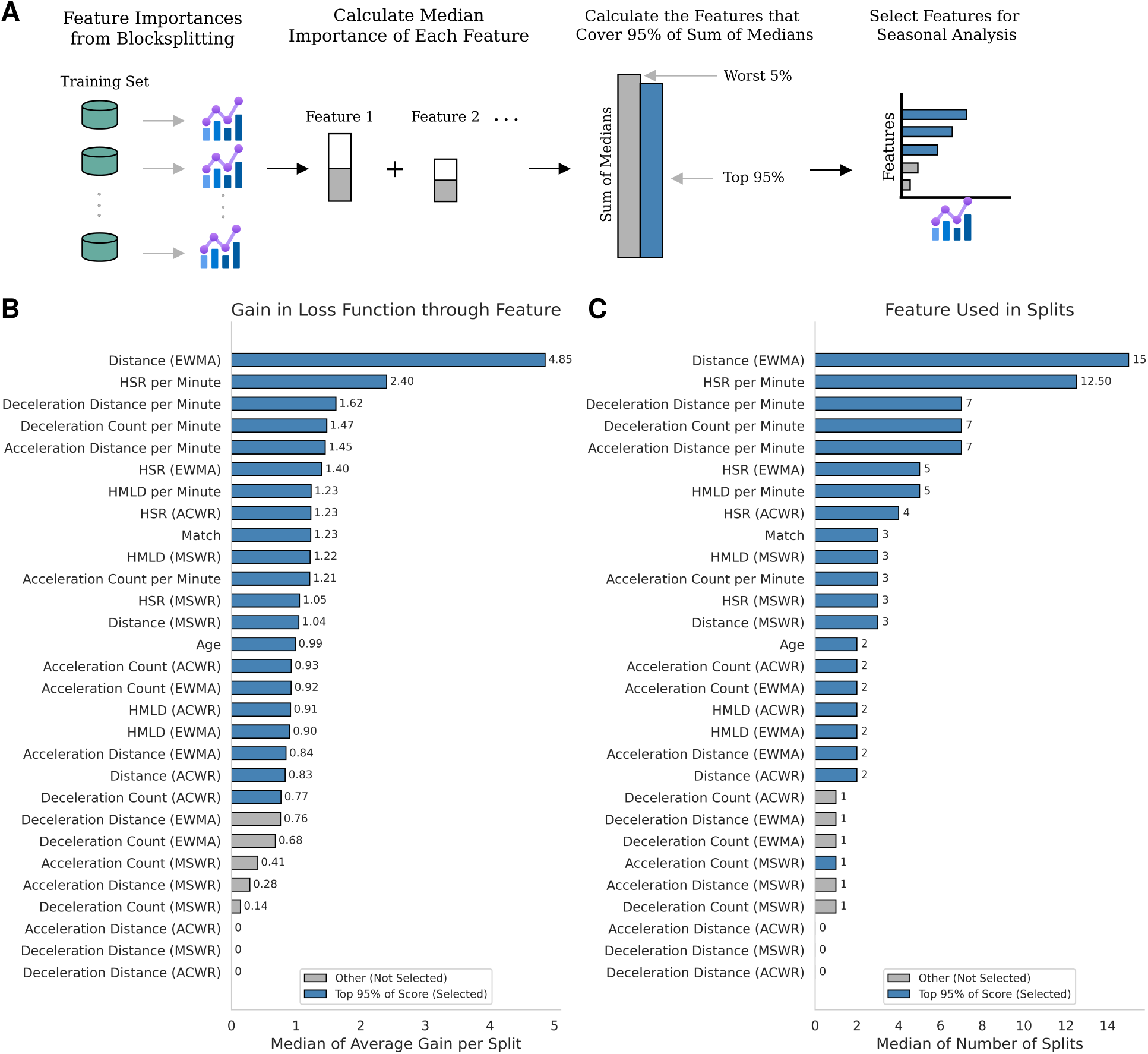
Feature Importance. Feature importances were computed for the XGBSE model using the feature importances of the underlying Accelerated Failure Time model. Inter alia, Exponentially Weighted Moving Averages (EWMA), Acute Chronic Workload Ratio (ACWR), and Monotonic Workload Ratio (MWR) were used. **(A)** The average gain in the loss function when selecting a feature and the number of times a feature is used in a split were calculated. The median of both metrics was computed across the 400 splits. Features with the lowest medians that collectively contribute less than 95% of the total sum of medians for both metrics are identified as uninformative. **(B)** Median of the average gain per split. Features contributing to 95% of the total sum of medians are highlighted in blue. **(C)** Median of the number of times a feature is used in a split within the tree ensemble. Features contributing to 95% of the total sum of medians are highlighted in blue.

The strongest predictor for both metrics was the exponentially weighted moving average of distance covered (in meters) over the past 21 days, capturing the cumulative effects of player load from prior playing activities. Following this, high-speed running intensity (HSR per minute) and the frequency of accelerations and decelerations on the day of activity emerged as the next most influential factors. In contrast, the cumulative acceleration and deceleration workload over the past 21 days had minimal impact, suggesting that while acute accelerations and decelerations on the day of activity contribute to injury risk, their accumulated effect over time plays a comparatively minor role in fatigue-induced injury susceptibility.

### Seasonal Prediction Validates Framework Performance with Positive Availability Gains

To evaluate the real-world applicability of our proposed framework in daily coaching scenarios, we applied the framework using the XGBSE model to predict player outcomes for the previously unseen 2022/2023 season. Predictions were generated in a weekly rolling fashion to simulate a realistic scenario where the model is re-trained weekly, incorporating the most recent data. This approach ensures that predictions are made using the latest trends and benefits from increasing training and calibration set sizes over time (Figure 7A). After obtaining all predictions, we aggregated the predicted labels into a test set for evaluating seasonal performance.

**Figure 7:**
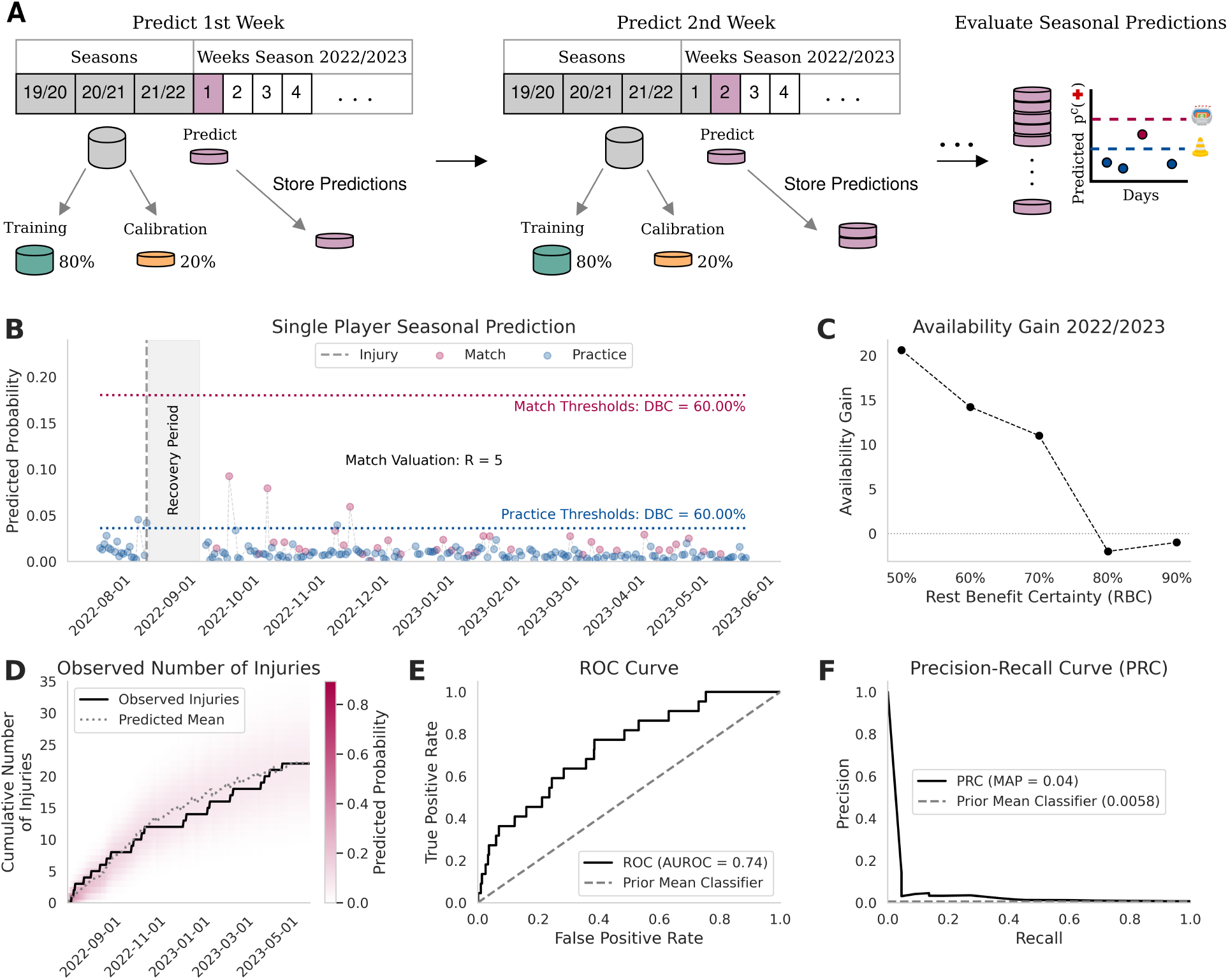
Predicting the 2022/2023 Season. **(A)** The previously unseen 2022/2023 season is predicted in a rolling weekly fashion. Seasons 2019–2022 are used as the baseline training set. Every 7 days, the model is re-trained using the baseline data and all available data up to that day, forming a training super set. Predictions for the subsequent week are made with the re-trained model. The training super set is randomly split into a training set and a calibration set, with 20% allocated to calibration. Weekly predictions are aggregated to evaluate the overall seasonal metrics. **(B)** Single-player results obtained with a match valuation of *R* = 5 and a Rest Benefit Certainty (RBC) of 60%. **(C)** Availability gain as a function of differing RBC values, evaluated on the accumulated test set. **(D)** Predicted distribution of the cumulative number of injuries per day (red) with its mean compared to observed injuries. The distribution is computed via iterative convolution over days. **(E, F)** Receiver Operating Characteristic (ROC) curve and Precision-Recall (PR) curve evaluated on the test set.

The features used for these predictions were selected based on the feature importance analysis of data from the 2019/2020 to 2021/2022 seasons (Figure 6A). To identify key predictors, we accumulated the medians for the gain in the loss function and selected features accounting for 95% of the cumulative importance.

To showcase how a seasonal trajectory for one player looks like, we show predictions for one player (Figure 7B) for a match valuation of *R* = 5 and a RBC of 60%, that highlight the potential of the framework. In August 2022, the player suffered one injury, which was correctly predicted by the model. After recovering from the injury, the player participated in three matches that exceeded the practice threshold but not the match threshold without sustaining further injuries. Hence, the model correctly classified these events as non-rest days. In November 2022, the model classified another rest day that did not result in an injury; this prediction was close to the classification threshold. Overall, the player would have been rested three times during the season: once when an injury occurred, once two days before an injury, and once when no injury occurred but the predicted injury probability was close to the classification threshold.

The availability gain curve for the 2022/2023 season, starting at a RBC of 50%, followed a decreasing trend, consistent with the split-based results from the 2019/2020 to 2021/2022 seasons (Figure 7C). Notably, RBCs of 50% and 60% yielded availability gains of around 20.1 and 14.7 match-weighted days, respectively, while RBCs of 80% and 90% resulted in slightly negative availability gains of −2.4 and −1.2 match-weighted days. These findings suggest that higher RBC thresholds, which require higher probabilities to classify a player as requiring rest (Supplementary Figure S4), may fail to prevent major injuries.

To evaluate the detection of injuries and the rate of misclassifying non - injury days as injuries, we further analyzed factors contributing to availability gain. This analysis included unnecessary rest days (false positives), saved days (prevented rest days due to detected injuries), as well as key performance metrics such as precision and recall (see Supplementary Figure S5). Across all match RBCs, the model consistently avoided predicting rest days during matches, demonstrating stable behavior. At a RBC threshold of 50%, the model identified 5 out of 22 injuries (recall: 22.73%), while only 1 in 18 predicted rest days corresponded to an actual injury (precision: 5.44%).

Results for match valuations *R* = 2, 10 were qualitatively the same and are reported within the Supplementary Figure S6 and Figure S7.

### Seasonal Predictions Align with Observed Data

To evaluate how well the predicted probabilities align with observed injury data, we computed the probability mass function of the cumulative number of injuries for each day. Our results demonstrate that the observed data aligns well with the high-probability mass regions predicted by the model (Figure 7D). The predicted mean trajectory closely follows the trajectory of the observed number of injuries. This implies that the model is well calibrated.

We further assessed model performance using the AUROC metric, achieving a score of 0.74 (Figure 7E). This value indicates a strong ability to discriminate between injury and non-injury events in the context of the high class imbalance with only 0.58% positive cases during the 2022/2023 season. Precision-recall analysis showed a Mean Average Precision (MAP) of 0.04, representing an approximately 6.9-fold improvement over random guessing (Figure 7F).

## Discussion

This study presented a novel injury prediction framework that integrates survival-based machine learning, probability calibration, and actionable decision thresholds tailored to day-specific valuations. These actionable insights are derived using rest-or-play thresholds derived from statistical decision theory. Therefore, predicted probabilities must accurately reflect actual injury risks. Beta calibration significantly enhanced the reliability of predicted probabilities, ensuring alignment with true injury risks and enabling actionable decisions.

Using a unique dataset from FC Barcelona’s first women’s team, we demonstrated that our framework, leveraging survival-based approaches, significantly outperforms standard machine learning classifiers^26–28^ in their ability to predict injury risk. The survival-based models can account for risk accumulation over time - a critical limitation of standard classifiers that are used in the literature^6;13;29^. Positive player availability gains - a metric, to the best of our knowledge, not previously reported in the literature - were observed across a range of match valuations and decision certainty thresholds, underscoring the framework’s robustness and flexibility in balancing injury prevention with team needs. The XGBSE model emerged as the top-performing method, validated through evaluation on three seasons of historical data and further applied to previously unseen data from the 2022/23 season, where it led to meaningful gains in player availability and closely aligned with the observed injury distribution.

We examined the most predictive features and identified fatigue-related measures, such as accumulated training distances of previous days, and same-day activity intensity emerged as the most influential predictors, showing that fatigue-related injury risk can be primarily predicted by previously long training sessions. Similarly, same-day intensity-related measures like acceleration and deceleration per minute account for a higher injury risk on that day.

The framework is highly flexible and, although demonstrated on professional soccer, is broadly applicable to other sports and domains. With the proliferation of wearable tracking devices not only among professional athletes but also amateurs and workers in physically demanding fields, our method offers a powerful tool for injury prediction in a variety of settings. The survival modeling approach seamlessly integrates features relevant to diverse injury mechanisms and domain-specific risk factors—ranging from biomechanical loads and psychological factors to specialized activity metrics. Even image or video data could straight-forwardly be incorporated to detect anomalies in movement patterns.

For other sport teams, we recommend re-training the model to capture differences in training regimes and player profiles. The computational efficiency of the XGBSE model - training on the entire dataset takes less than a second (Supplementary Figure S9) - makes re-training feasible even for large datasets.

Despite its strengths, our method has limitations. First, similar to standard evaluation metrics like the F1 score or balanced accuracy, the availability gain metric does not represent a causal intervention effect. Specifically, (a) our evaluation does not fully account for scenarios where resting a player before an injury could have prevented it, as shown in the seasonal analysis, and (b) resting a player on a high-risk day may prevent an injury that could still occur later. Since (a) underestimates and (b) overestimates the real-world effect, the net impact remains unclear, precluding causal interpretation. Notably, while this limitation applies to all metrics, availability gain uniquely accounts for both injury occurrence and injury severity. Second, while our dataset from FC Barcelona’s women’s team spans four seasons, validation on data from other clubs would enhance generalizability. Differences in training, playing styles, and player demographics could provide further insights into the framework’s robustness.

Future improvements could enhance both the methodological and practical aspects of the framework. Its fast computation enables real-time applications, allowing the integration of live match or training data for immediate insights and rapid, data-driven decisions - potentially even during matches or training. A long-term goal is the development of digital twins that simulate player states, team dynamics, and causal injury intervention effects in real time. By integrating our model within multiple modeling approaches - such as match simulations - these digital twins could optimize data-driven decision-making for teams.

Incorporating player-specific metrics, such as salary or market value loss, into the loss function could help prioritize long-term injury prevention. Additionally, incorporating informative features - such as heart rate data - and employing random effect models or Gaussian processes to capture temporal correlations in survival curves may further enhance personalized predictions.

Overall, this work demonstrates the capability and practicality of using survival-based machine learning combined with probability calibration and statistical decision theory to predict injury risks and inform actionable decisions in elite sports. By incorporating day-specific valuations and decision-maker certainty thresholds, our approach bridges the gap between academic research and real-world deployment. Its scalability, flexibility, and transferability make it a valuable tool for sports organizations seeking to optimize player availability, performance, and long-term outcomes across diverse settings.

## Methods

### Employed Models

To predict injury risk in professional footballers, we employ both parametric and non-parametric survival models. Parametric models, such as the penalized Cox proportional hazards model (Coxnet)^18;33;34^, specify a functional form for the hazard rate, offering well-understood statistical properties. Non-parametric models, including the Extreme Gradient Boosting Accelerated Failure Time model ^17;35^ and the boosted Cox model^30;34^, provide the flexibility to model complex, non-linear dependencies but lack the parametric guarantees of convergence.

To explore the predictive power of combining machine learning classifiers as feature embedding tools, we use the Extreme Gradient Boosting Survival Embedding (XGBSE) model ^16^, which combines an Accelerated Failure Time framework as tree-based embedding with logistic regressions to estimate bin-level survival probabilities^36^.

As a control, we also evaluate predictions using classification models, including LightGBM^26^, LightGBM with monotone risk increase for activity duration, logistic regression with Light-GBM tree embeddings, k-Nearest Neighbors^27;37^, and logistic regression^28;37^. All classification models incorporate Bayesian hyperparameter optimization^38^.

### Calculating Injury Probabilities Based on Survival Curves

All surival-based approaches estimate a survival function *S*_*θ*_ : (0, *∞*] *→* [0, 1], where the parameterization *θ* depends on the specific method employed. In Cox-based models, such as Coxnet^18^, 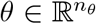 represents the coefficients of the proportional hazards model. Tree-based methods, including Accelerated Failure Time^17^ and Boosted-Cox^30^, parameterize *θ ∈* 𝔽, where 𝔽 denotes the space of additive tree ensembles. The XGBSE model^16^ combines these approaches, with 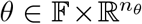, comprising a tree ensemble for the underlying Accelerated Failure Time model and real-valued parameters for logistic regressions used in post-processing.

Given a parameterization *θ*, the survival function *S*_*θ*_(*m*|*x*), conditioned on covariates 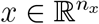, takes as input the number of player minutes *m* and outputs the probability that an individual does not sustain an injury up to minute *m*. The probability of sustaining an injury up to minute *m* is expressed as ℙ_*θ*_(*m*|*x*) = 1 *− S*_*θ*_(*m*|*x*).

The monotonicity of the survival function ensures that the probability of injury increases as the number of minutes played increases. As our data reports only the actual minutes played, which serve as lower bounds for planned activity durations on injury days, we compute *p*_*i*,*t*_ on injury days as the mean injury probability for all training durations of players who trained at least as long as the injured player on that day. For non-injury observations, we use the recorded time of activity.

Given that readers may not be familiar with the XGBSE model, we provide a detailed explanation of its approach to estimating survival curves in the supplementary material.

### Probability Calibration with Beta Calibration

To adjust the estimated probabilities to align with observed injury frequencies, we employed probability calibration for all models. Using the model parameters 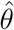 estimated from the training set *𝒟*^Train^, we calculated the uncalibrated probabilities 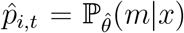 for each observation in the calibration set *𝒟*^Cal^, consisting of unseen data points (*y*_*i*,*t*_, *x*_*i*,*t*_, *m*_*i*,*t*_).

The uncalibrated probabilities 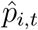 were adjusted using beta calibration ^19^, which applies the following mapping function:

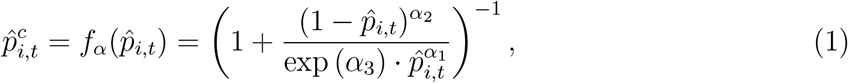

where *α* = (*α*_1_, *α*_2_, *α*_3_)^*′*^ are the parameters of the calibration function. These parameters were estimated by fitting the calibration function to the calibration set *𝒟*^Cal^.

### Day Importance Weighting

To account for the varying importance of a player missing a particular day, we introduced the concept of match importance. Match importance assigns an individual cost *c*_*i*,*t*_ *∈* ℝ_+_ to each day *t* for a player *i*, reflecting the impact of their absence.

These costs are specified by decision-makers and can be dynamically adjusted to reflect context-specific priorities, such as the higher importance of cup knockout matches compared to practice sessions. As the definition of these costs is highly club-specific, we distinguished for our analysis between the cost of missing a match *c*^(*G*)^ *∈* ℝ_+_ and the cost of missing a practice *c*^(*P*)^ *∈* ℝ_+_, both of which are assumed to be constant across individuals and days.

Since only the relative values of *c*^(*G*)^ and *c*^(*P*)^ are relevant for our analysis, we defined the relative match importance ratio *R* = *c*^(*G*)^*/c*^(*P*)^. This ratio is used to transform practice days into match-weighted practice days by dividing the number of practice days by *R*. In an application of a club the ratio *R* can be day and individual specific, allowing individualized tailored weighting of days for players and competitions.

### Cost of Playing and Resting

The decision for a player to participate in a match or practice session carries a potential cost, depending on whether the player sustains an injury. To quantify this, we defined a cost function incorporating the predicted injury label *ŷ*_*i*,*t*_ *∈* {0, 1}, the actual injury outcome *y*_*i*,*t*_ *∈ {*0, 1*}*, the number of missed matches 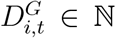 and practices 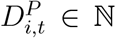, the match indicator *G*_*i*,*t*_ (1 for match days, 0 otherwise), and the match importance ratio *R*.

If a player rests, the cost reflects the missed day, with a value of 1 for match days and 1*/R* for practice days. If a player participates and sustains an injury, the cost accounts for the missed matches and practices, expressed in match-weighted days. To consolidate these cases, we defined the overall cost function

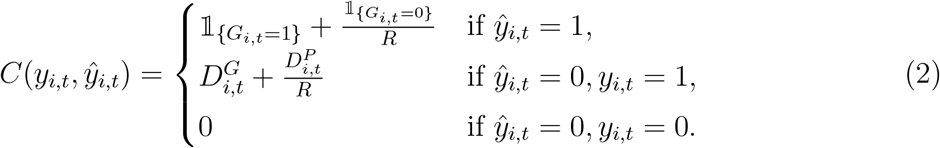

This cost function provides a unified framework for quantifying decision outcomes and facilitates the calculation of conditionally expected costs, discussed in the next subsection.

### Conditionally Expected Costs Under Injury Duration Uncertainty

The cost function *C*(*y*_*i*,*t*_, *ŷ*_*i*,*t*_) depends on the actual injury outcome *y*_*i*,*t*_, the number of missed matches 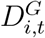, and missed practices 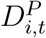 in the case of an injury. However, a priori, the injury outcome and injury durations are unknown.

Using the estimated calibrated probabilities, the expectation of the costs, conditioned on 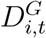 and 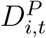, is dependent on the decision to rest a player (*ŷ*_*i*,*t*_) and is expressed as:

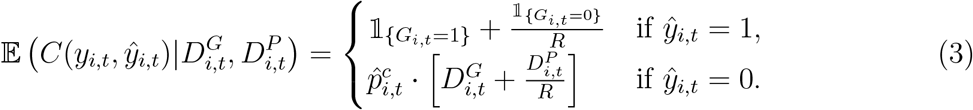

If *ŷ*_*i*,*t*_ = 1, the conditionally expected costs incorporate stochasticity due to the uncertainty in the injury outcome. The player incurs a cost associated with an injury, weighted by the calibrated injury probability 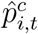, or no cost if no injury occurs with probability 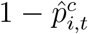. If *ŷ*_*i*,*t*_ = 0, the costs are deterministic because the player rests and misses the session, and the conditionally expected costs are equal to the cost function for resting.

Since 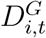 and 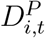 are unknown a priori, the conditionally expected cost 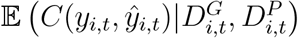 is itself a random variable that depends on the underlying distribution of 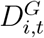 and 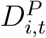.

### Augmenting Data Set with Historical Injury Durations

The distribution of missed matches and practices due to injury is critical for deriving the distribution of conditionally expected costs. To estimate this, we utilized historical data and augmented it with additional data from Soccerdonna^39^.

Our dataset initially contained 83 recorded non-contact injuries. To improve the robustness of the injury duration distribution estimate, we included injury data from players listed on the first teams of the top female football divisions in Spain (Primera División de la Liga de Fútbol Femenino), Germany (Frauen-Bundesliga), England (Women’s Super League), and the United States (National Women’s Soccer League) during the 2022/2023 season. This augmented dataset comprised injury durations *D*_*j*_ for *j* = 1, 2, ߪ, 494, significantly increasing the sample size by 411 observations. We included all injuries indicative of non-contact mechanisms; a complete list of the selection criteria is provided in the Supplementary Table S1.

To estimate the number of missed matches and practices from total injury durations, we applied a linear regression approach to partition the injury durations into match days and practice days. Specifically, we fitted the following regression, once for both *k* = *G, P*, on the combined training and calibration:

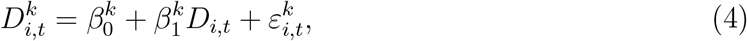

where *D*_*i*,*t*_ is the total injury duration recorded for individual *i* on day *t*. The fitted parameters 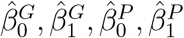 were then applied to the augmented injury dataset to estimate the number of missed matches 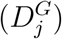 and practices 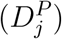 for each injury observation. The distribution of *D*_*j*_, as well as the fitted linear regression lines for the entire dataset, are provided within the Supplementary Figure S8.

### Decision Thresholds and Rest Benefit Certainty

Given the calibrated probabilities 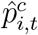, the cost function *C*(*y*_*i*,*t*_, *ŷ*_*i*,*t*_), and the joint distribution of missed matches 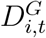 and practices 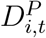, decision-makers must determine whether a player *i* should play or rest on day *t*. This decision is based on whether the expected cost of playing exceeds the expected cost of resting:

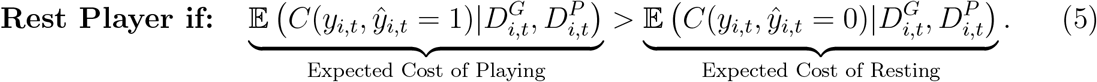

Using the previously derived formula for expected costs, this inequality can be reformulated in terms of the calibrated probabilities 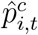, the match indicator 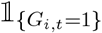, and the match valuation parameter *R*. Solving for 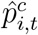 provides the following decision thresholds

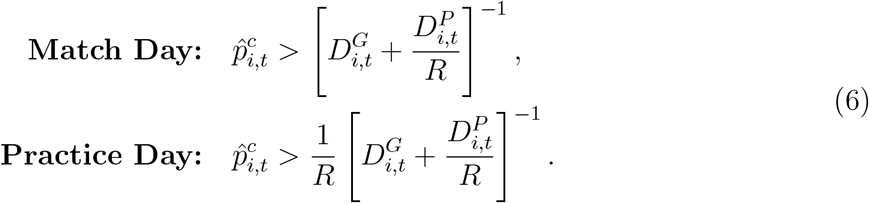

For match days, it is optimal to rest a player if the probability of injury exceeds the inverse of the match-weighted injury duration. For practice days, this threshold is further adjusted by the inverse match valuation parameter *R*. This adjustment reflects that, all else being equal, the probability of injury must be *R* times higher on match days than on practice days to justify resting the player.

Using the joint distribution of 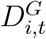 and 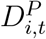, the probability *Q*_*i*,*t*_ that the threshold condition in Inequality 6 is satisfied can be computed. For match days, this probability is given by:

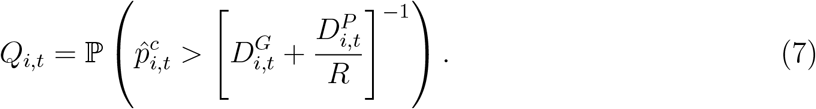

The value *Q*_*i*,*t*_ represents the probability that resting a player is the optimal decision. Decision-makers specify a threshold level of certainty, referred to as the Rest Benefit Certainty (RBC). If *Q*_*i*,*t*_ *>* RBC, the player is rested; otherwise, she plays.

### Availability Gain Evaluation Metric

The Availability Gain (AG) metric evaluates decision-making strategies by balancing the negative impact of unnecessary rests with the positive impact of detecting injuries that result in missed match and practice days. Days are weighted as match days using the match valuation parameter *R*.

The metric assigns a negative value to rested days, regardless of whether an injury occurs, and a positive value to detected injuries. It is defined as:

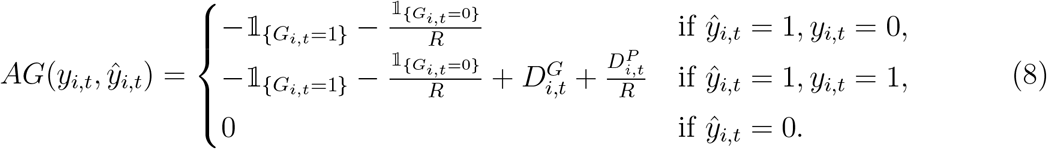

In this formulation, the negative term corresponds to the cost of resting, and the positive term accounts for the benefit of detecting injuries in terms of missed match 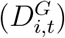 and practice days 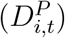. The AG metric over the test set is computed as the sum of availability gains across all test set observations.

### Interpretation of Mean Average Precision and Area Under the Receiving Operator Curve

We quantified discrimination using the Mean Average Precision (MAP) and the Area Under the Receiver Operating Characteristic Curve (AUROC). MAP evaluates how well a model balances precision and recall, making it particularly suited for imbalanced datasets because it penalizes false positives and rewards correct identification of the minority class. In contrast, AUROC represents the average sensitivity and specificity across varying decision thresholds, providing a broader assessment of performance.

### Injury Data Recording

The medical team managed and documented injuries using a validated electronic medical record system (COR version 2.0; FCB). Injury assessments were performed by the team’s medical physician, in conjunction with the FC Barcelona medical department, following standardized diagnosis and return-to-play protocols as outlined in the club’s guidelines ^24^. The analysis focused on non-contact injuries affecting muscles, tendons, ligaments, and cartilage.

### Recording of Workload Features

The external workload during training sessions and matches was monitored using GPS data obtained from the WIMU PRO™ device (RealtrackSystems S.L., Almeria, Spain)^40^. The collected data underwent pre-processing through SPRO™ Software (version 927) (RealtrackSystems, Almeria, Spain). This software compiled the data in RAW format and facilitated the generation of continuous training-load variables. The GPS training-load variables used in this study for include the distance covered in meters and the high metabolic load distance (HMLD; measured in meters, defined as the distance covered when metabolic power exceeds 25.5 Watt/kg, at speeds above 5.5 m/s). Additionally, the number of accelerations (higher than 3 m/s2) and decelerations, along with the respective distances in meters covered during accelerations as well as decelerations, are considered. Finally, the distance covered while high-speed running per minute (relative HSR) is included, defined as running at speeds above 18 km/h.

### Feature Engineering

Baseline features included per-minute training load variables derived from player tracking data. For the classification comparison models, the duration in minutes was added as an additional feature. To capture training load dynamics, new variables were designed following the methodology of Rossi et al.^6^. These included exponentially weighted moving averages, with 21 features calculated to represent past training loads. The acute chronic workload ratio was defined as the ratio of the load over the last six days to the load over the last 28 days, assessing shifts between acute and chronic training loads. The monotonic workload ratio was calculated as the mean load over the last seven days divided by the standard deviation during the same period, capturing variability in recent training loads. Additional features incorporated into the models were age, treated as a continuous variable, and a binary match indicator variable.

The list of all used features and their computations, if applicable, are given within the supplementary Table S3.

### Feature Pre-Processing

For numerical features used in the Coxnet, k-Nearest Neighbors, and Logistic Regression models, standardization was performed using the mean and standard deviation of each feature. Principal Component Analysis (PCA) was then applied to reduce dimensionality, retaining components that captured 95% of the total variance in the data.

### Hyperparameter Optimization for Classification Models

Hyperparameter optimization for the standard machine learning models was carried out using Bayesian hyperparameter optimization ^38;41^ with 60 iterations, which is standard in the literature^42^. The final parameters and details on the hyperparameter search space are provided in the Supplementary Material.

## Implementation and Code Availability

All models were implemented in Python 3.11.8. The survival-based models were implemented as follows: the XGBSE model utilized the xgbse package version 0.3.3, the Accelerated Failure Time model was implemented with the xgboost package version 2.1.1, and the Coxnet and CoxBoost models were developed using the scikit-survival package version 0.23.0. The classification comparison models, including LightGBM, LightGBM with monotone duration of activity, k-Nearest Neighbors, and Logistic Regression, were developed using scikit-learn version 1.5.2. The LightGBM Tree Embeding model was implemented using the same Light-GBM and Logistic Regression setup. Code will be made available from the corresponding author upon reasonable request.

## Data Availability

Data will be available upon reasonable request, considering ethical and privacy concerns, from the author Gil Rodas.

## Ethical Considerations

This study adhered to the Declaration of Helsinki guidelines ^43^ and received approval from both the Barça Innovation Hub’s local committee and the Ethics Committee of Consell Català de l’Esport (code 012/CEICGC/2021). Participants were informed about the study’s potential risks and benefits. All personal data and results were anonymized to maintain confidentiality and comply with the criteria specified by the General Data Protection Regulation (GDPR).

## Author Contributions

M.H. developed the statistical and mathematical theory and implemented the models in Python. B.C. conducted robustness checks. E.F., X. Y. and G.R. collected the data. M.H. visualized the results. M.H., J.H., and J.R.G. conceptualized the study. M.H., J.H., and J.R.G. wrote the manuscript. J.H. and J.R.G. oversaw the research process. All authors reviewed and approved the final version.

## Supplementary Material to

This supplementary material provides additional technical details, illustrative examples, and methodological insights that complement our main study, Informed Injury Prediction in Elite Football: Decision Theory meets Machine Learning. We begin by presenting detailed examples of individual decision-making, where we contrast the expected costs of resting versus playing under various injury scenarios and illustrate the concept of Rest Benefit Certainty (RBC). Next, we showcase the availability gain metric to quantify how different injury durations affect player availability, incorporating the impact of match valuation.

Further, we describe the Extreme Gradient Boosting Survival Embedding (XGBSE) model used for estimating survival curves from censored injury data, outlining its three-step procedure—from fitting a censored Accelerated Failure Time model to feature space transformation and logistic regression on binned survival times. Finally, we explain our imputation strategies for integrating national team training exposure into the overall training history, ensuring that gaps in the data are appropriately addressed.

### Illustrative Example: Individual Decision-Making

Consider a scenario where decision-makers must decide whether a player should rest or participate in training the next day. The model predicts a 5% probability of injury for the upcoming day, which is a practice day. Injuries are assumed to last, with equal probability, 7, 14, or 21 days. Matches occur once per week, and matches are valued *R* = 5 times more than practices.

#### Expected Cost of Resting vs. Playing

The cost of resting the player on a practice day is given by the match-weighted value of a practice day:

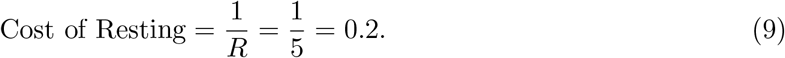

The expected cost of playing, conditioned on the injury duration, is calculated as follows:

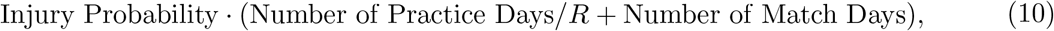

such that

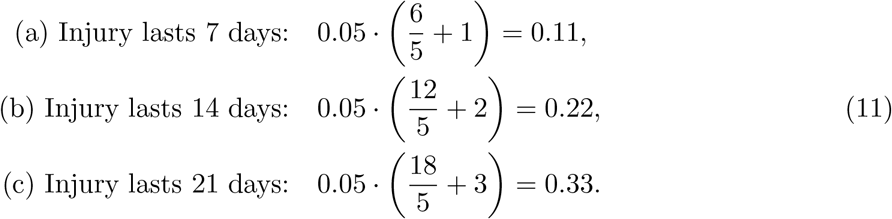

Comparing the resting cost of 0.2 to the conditional expected costs of playing, we observe that only in scenario (a), where the injury lasts 7 days, playing yields a lower cost. In 66.67% of cases (scenarios b and c), playing has a higher expected cost, making resting the favorable decision under these assumptions.

#### Rest Benefit Certainty (RBC)

Decision-makers may require a minimum level of certainty, referred to as Rest Benefit Certainty (RBC), to rest the player. For instance, if the required certainty level is 50%, resting is optimal because playing has a higher expected cost in 66.67% of cases. However, if the required certainty is 70%, decision-makers would not rest the player. The RBC represents the required confidence level to justify resting but does not directly affect the expected costs.

#### Influence of Match Valuation

The match valuation parameter *R* directly influences the costs. If matches are valued only 2 times more than practices (*R* = 2), the cost of resting becomes:

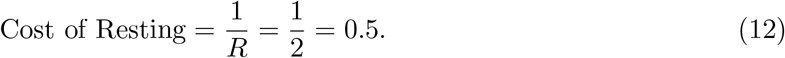

The expected costs of playing are recalculated as:

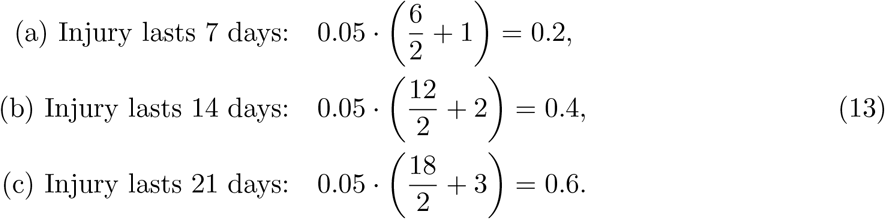

Comparing these values to the new resting cost of 0.5, we see that only in scenario (c), where the injury lasts 21 days, resting yields a lower cost. As this outcome has a probability of 33.33%, decision-makers with a RBC of 50% would now decide to let the player participate.

This example highlights how match valuation influences decision-making. When matches are highly valued (*R* = 5), even short injuries justify resting the player. Conversely, when matches are less valued (*R* = 2), only long injuries justify resting. The decision to rest or play a player critically depends on the day-specific valuation of matches versus practices.

### Illustrative Example: Availability Gain Metric for Varying Injury Durations

To illustrate the application of our proposed availability gain metric, consider a scenario with 15 observed injuries in the test set, where matches are valued 10 times more than practices (*R* = 10).

#### Scenario 1

The model correctly predicts one injury, preventing two missed practice days. However, the model also incorrectly predicts injuries on 50 other occasions, leading to 51 lost practice days (including the day of true prediction). The availability gain is calculated as:

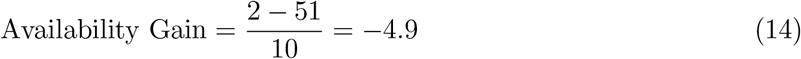

This results in a net loss equivalent to 4.9 match days, indicating that the cost of false positives outweighs the benefit of the correct prediction in this scenario.

#### Scenario 2

In contrast, if the correctly predicted injury prevents a 30-day absence, including 3 matches and 27 practices, then accounting for the false positives, the net benefit becomes:

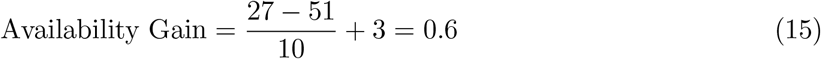

This represents a net gain in player availability, expressed in match-weighted valuation.

These examples highlight how injury duration significantly influences the player availability and how clubs can adjust *R* to reflect specific priorities, such as differentiating between practices, friendly matches and critical knockout games.

### Estimating the Survival Curve with the XGBSE Model

The Extreme Gradient Boosting Survival Embedding (XGBSE) model ^16^ is a three-step procedure for estimating survival curves from censored survival data. The method combines tree-based modeling with logistic regression to provide a robust framework for analyzing survival distributions. The three steps include: 1. Fitting a censored Accelerated Failure Time model on daily injury observations. 2. Transforming the feature space based on the fitted Accelerated Failure Time model. 3. Using logistic regression to estimate binned survival time probabilities in the transformed feature space to estimate the survival curve.

#### Step 1: Fitting the Accelerated Failure Time Model

The first step involves fitting the following Accelerated Failure Time model to the data:

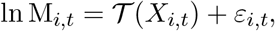

where M_*i*,*t*_ represents the number of minutes player *i* plays on day *t* until injury, *X*_*i*,*t*_ is the corresponding feature vector, *𝒯* (*X*) is a tree-ensemble model, and *ε*_*i*,*t*_ *~𝒩* (0, *σ*^2^) is Gaussian noise, conditionally independent and identically distributed. Injuries are rare events, and for days without injuries, the actual survival time is unknown. To handle this, a censored likelihood approach is employed.

The density of an observation, conditioned on the injury indicator *Y*_*i*,*t*_, is given by:

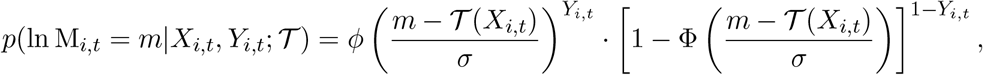

where *ϕ* and Φ denote the standard normal probability density and cumulative distribution functions, respectively. The tree ensemble *𝒯* is estimated by minimizing the negative log-likelihood:

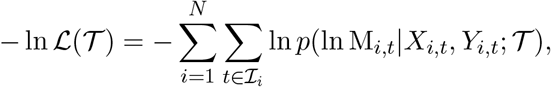

where *ℐ* _*i*_ is the set of all observations for individual *i*. This approach allows the model to handle right-censored data effectively, where exact injury times are unavailable.

#### Step 2: Feature Space Transformation

After fitting the Accelerated Failure Time model, the tree ensemble *𝒯* is used to transform the feature space. Each of the *K* trees in the ensemble has *n*_*k*_ leaf nodes, and each observation is assigned to exactly one node per tree. This creates a new high-dimensional, sparse feature space represented by indicator variables 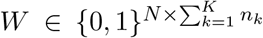. Each row of *W* contains *K* indicator variables set to one, corresponding to the leaf nodes an observation falls into, while the remaining entries are set to zero.

#### Step 3: Logistic Regression on Binned Survival Times

The transformed feature space *W* is then used to fit logistic regressions on binned survival time indicators. Observations are grouped into five disjoint time intervals, each spanning 30 minutes, denoted as [30 *·* (*r −* 1), 30 *· r*). For each interval *r*, the logistic regression estimates the conditional probability 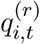 of injury occurring within that interval, given that the individual survived until its start.

The survival function is computed by combining these probabilities:

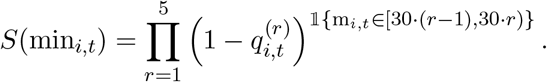

This formulation ensures that the model accurately accounts for survival probabilities over time, incorporating both censored and uncensored observations.

### Augmenting Activity Load Records by Imputing National Team Records

To address missing training load data, unreported days were classified as either rest days or missing values based on predefined rules (see Supplementary Table S2). For players on national duty, training load was imputed using a three-step process. First, the distance covered in national matches was predicted using a mixed-effects model that incorporated match minutes and player position. Second, training loads for national training days were estimated using a gradient boosting model with predictors such as time since the last match, time until the next match, and player position. Finally, additional training exposure variables were derived using another gradient boosting model, which used the imputed distances as inputs and accounted for player-specific variability.

## Supplementary Figures

**Figure S1:**
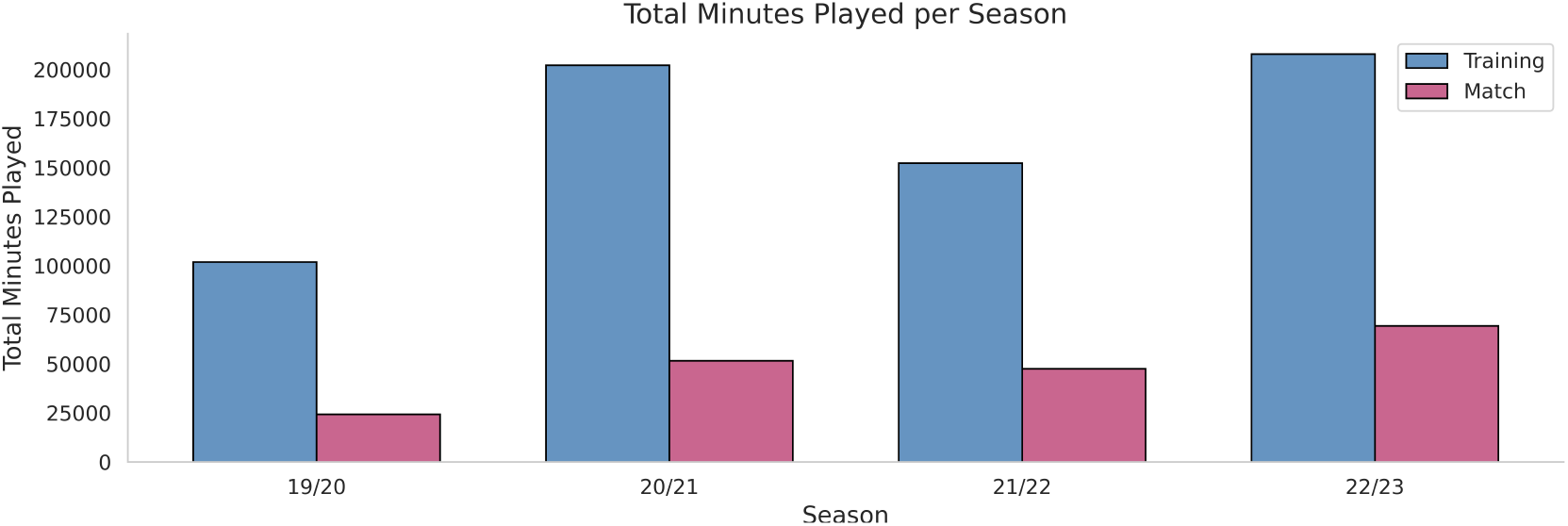
Played Match and Practice Minutes. The total time played in minutes per season.

**Figure S2:**
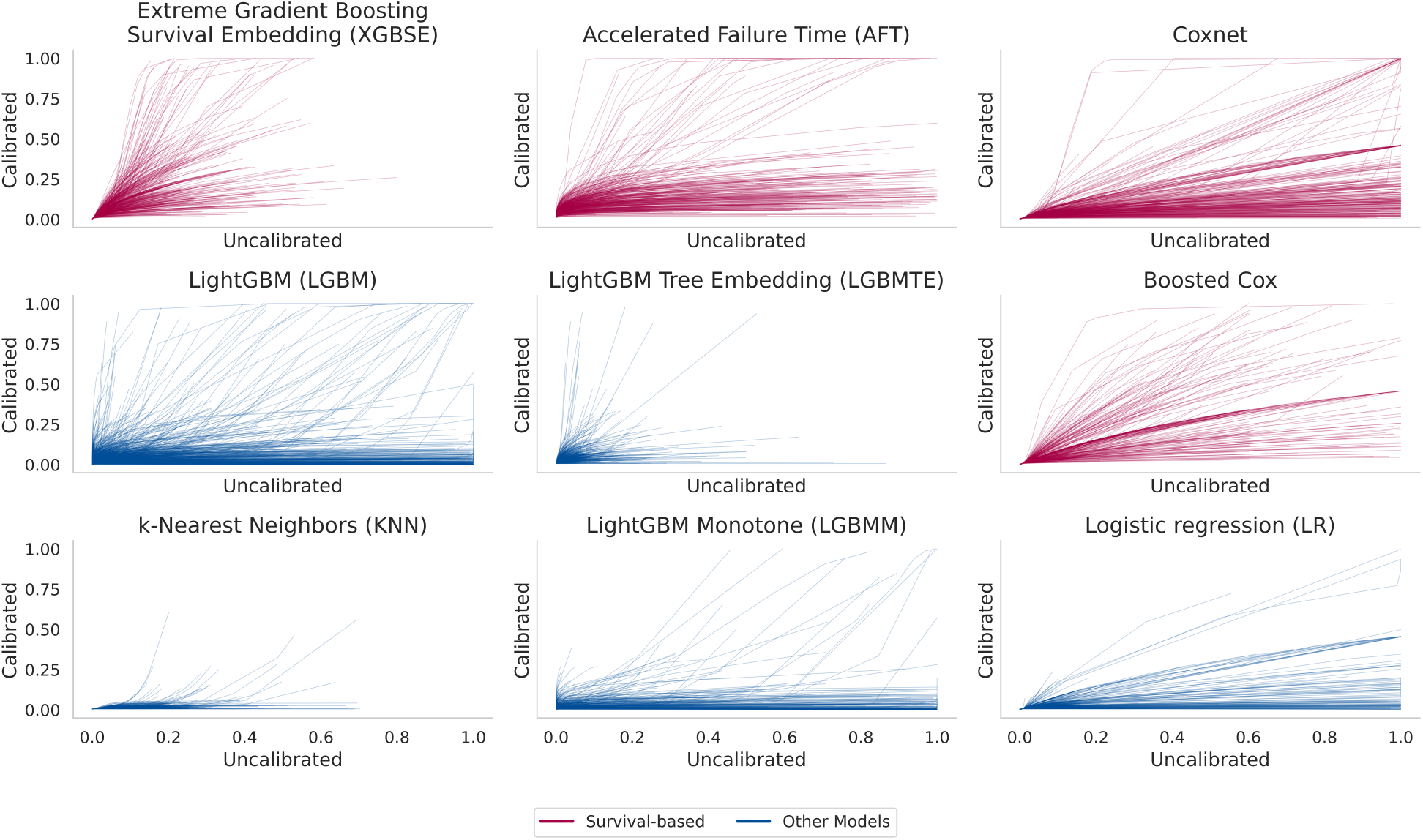
Beta Calibration Transformations Over Sample Splits. Each curve represents one fitted beta calibration function on the validation set. One curve is calculated for each of the 400 block splits. Survival-based models are given in red and standard classification models in blue.

**Figure S3:**
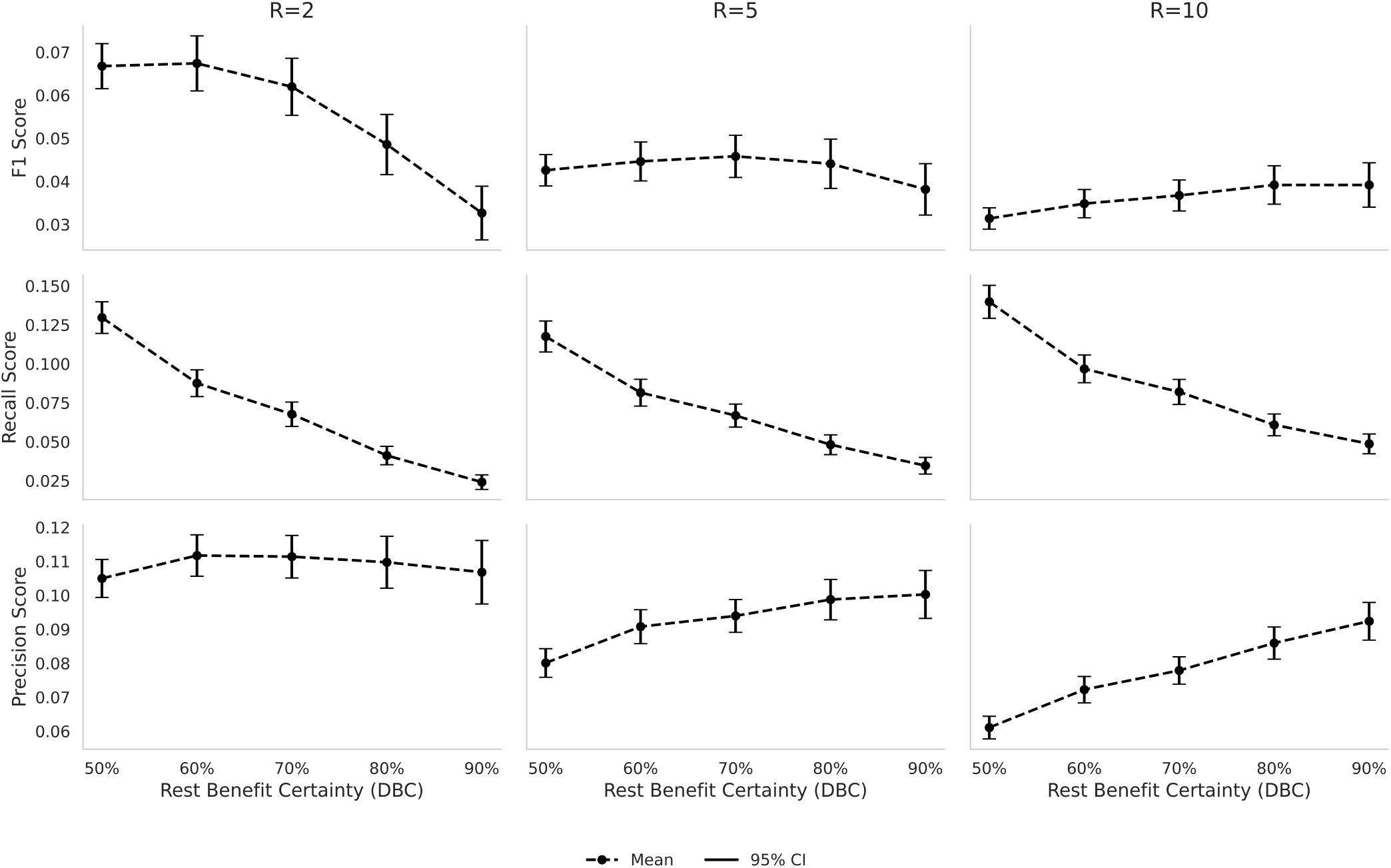
Mean F1-Score, Precision, and Recall Over Sample Splits. Each point represents the mean of the respective metric evaluated at the respective Rest Benefit Certainty (RBC) over the 400 sample splits. Confidence intervals represent the 95% confidence intervals around the respective means.

**Figure S4:**
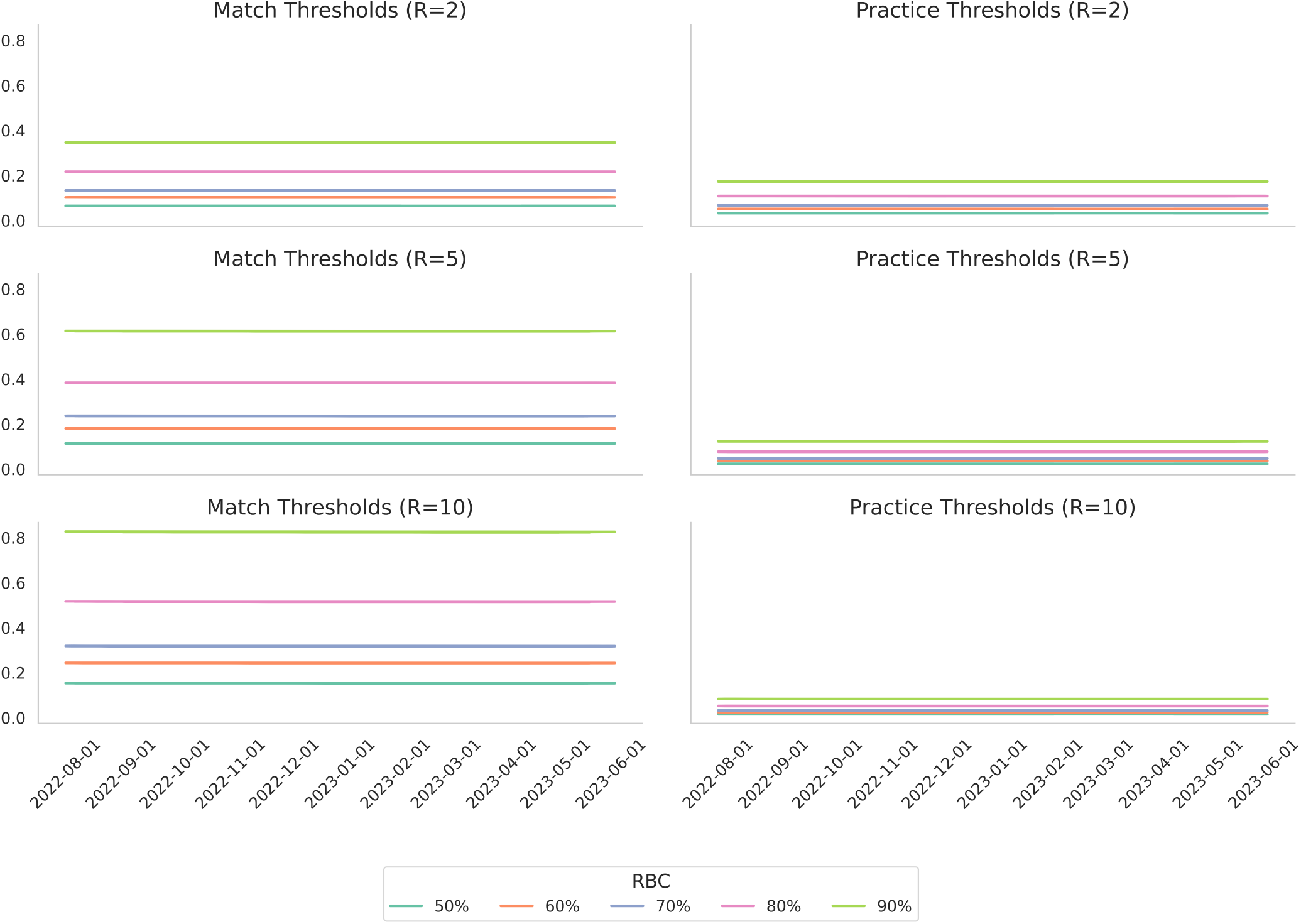
Decision Thresholds According to Match Valuation and Type of Activity for the 2022/2023 Season. Each line represents a match or practice threshold, depending on the Rest Benefit Certainty (RBC) and match valuation (*R*) for the 2022/2023 season. By construction of the RBC, the order of the lines is the same across plots, only the magnitudes change.

**Figure S5:**
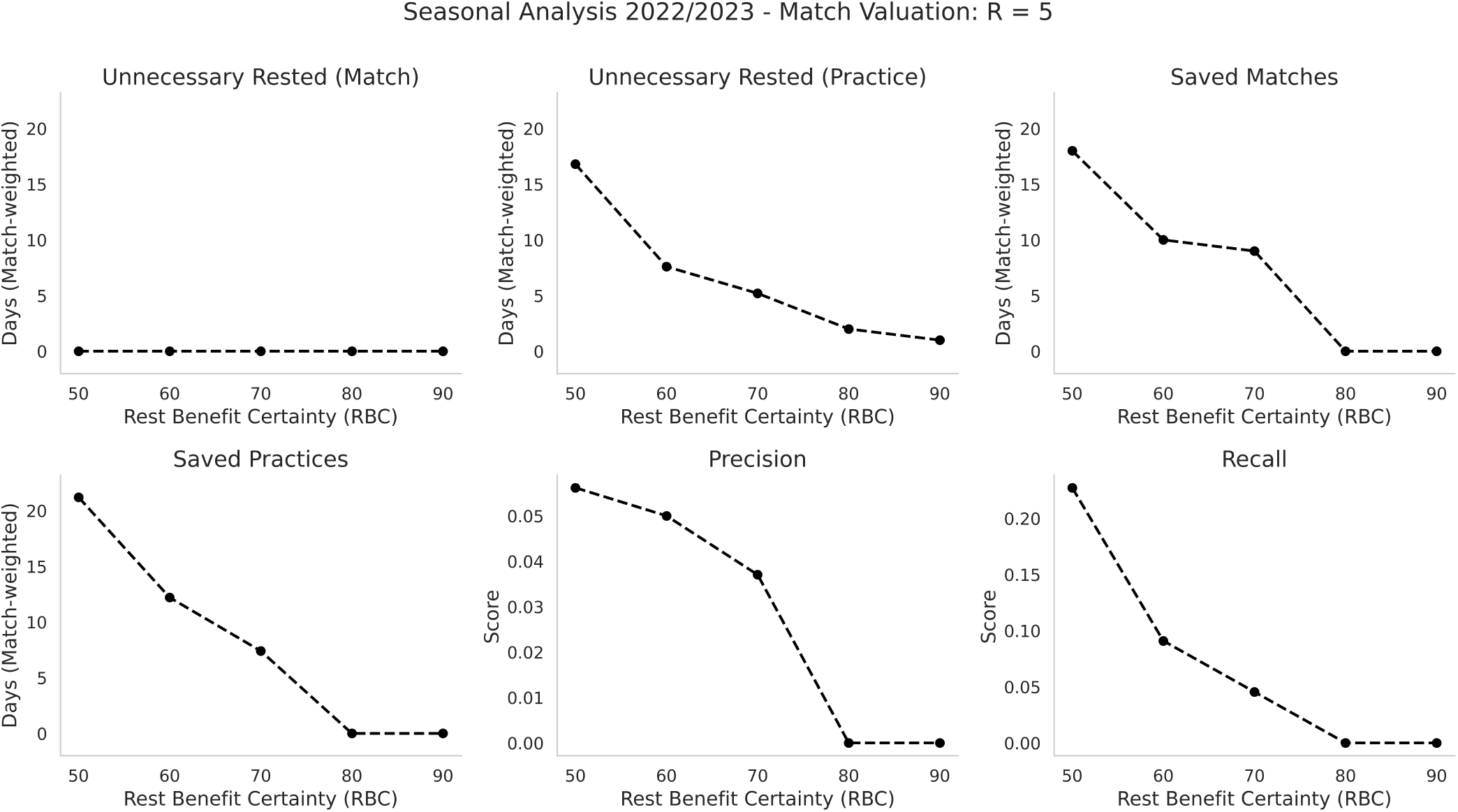
Additional Evaluation Metrics for Predicting the 2022/2023 Season with R = 5. Each point represents the respective metric evaluated at the respective Rest Benefit Certainty (RBC) for the 2022/2023 season.

**Figure S6:**
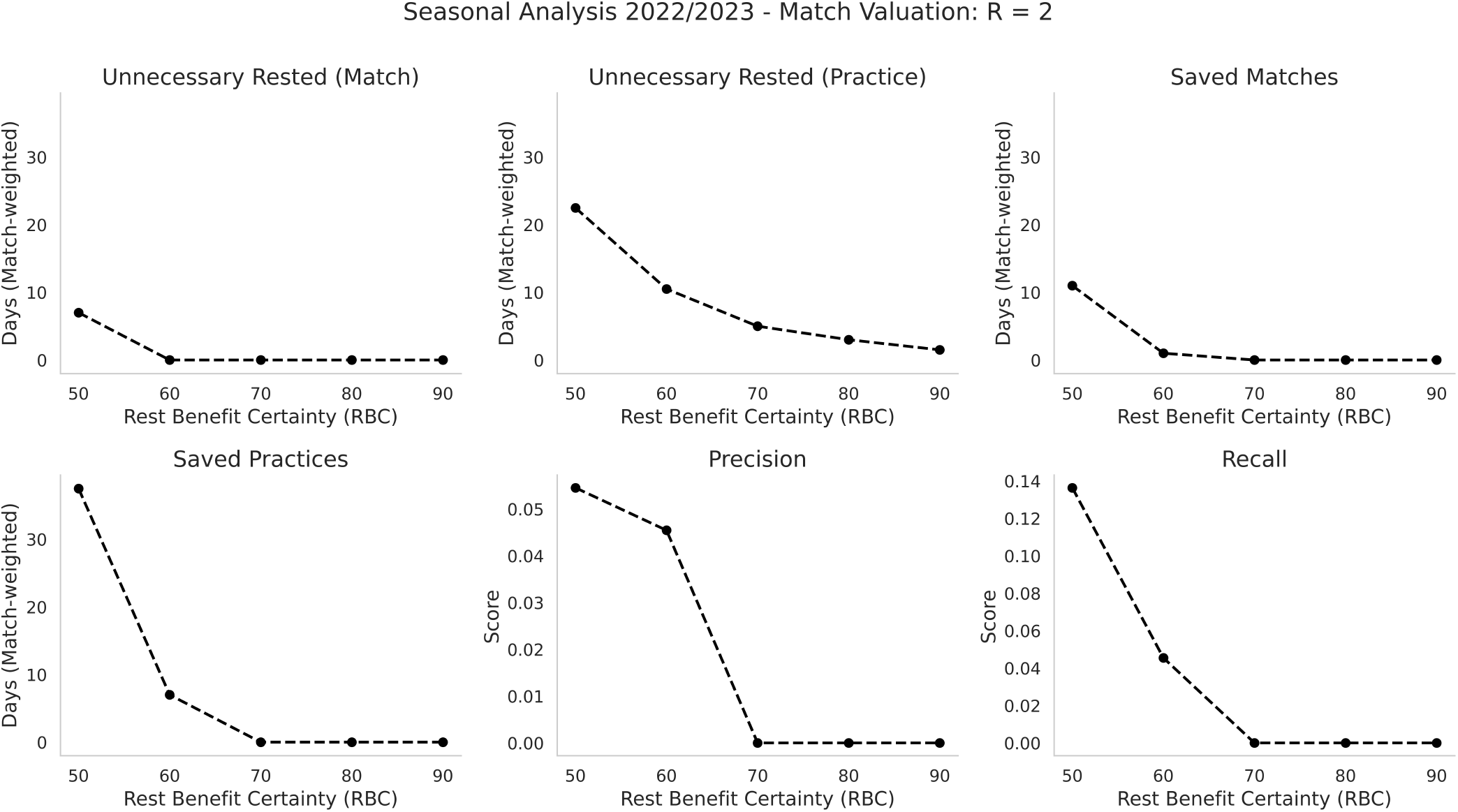
Additional Evaluation Metrics for Predicting the 2022/2023 Season with R = 2. Each point represents the respective metric evaluated at the respective Rest Benefit Certainty (RBC) for the 2022/2023 season.

**Figure S7:**
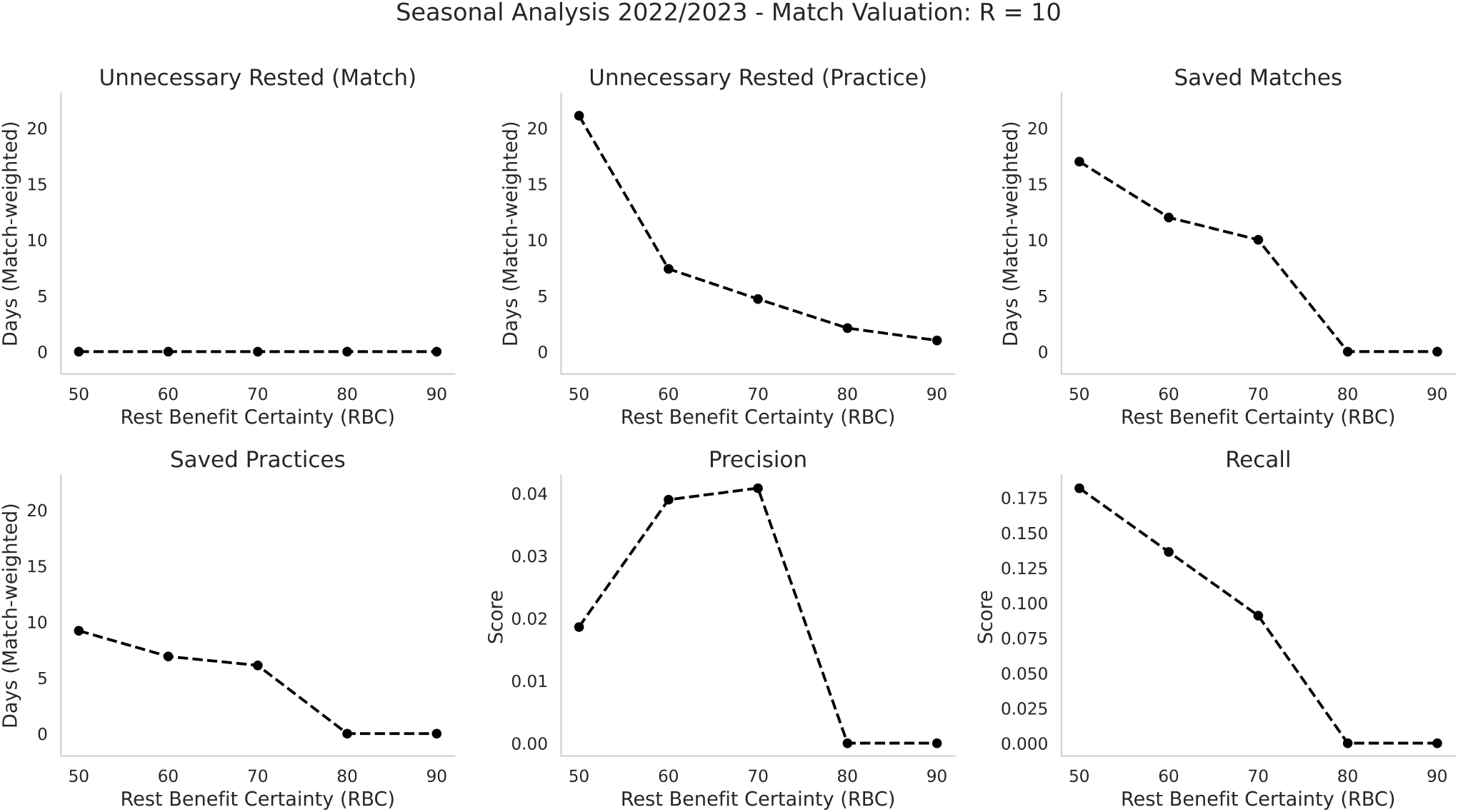
Additional Evaluation Metrics for Predicting the 2022/2023 Season with R = 10. Each point represents the respective metric evaluated at the respective Rest Benefit Certainty (RBC) for the 2022/2023 season.

**Figure S8:**
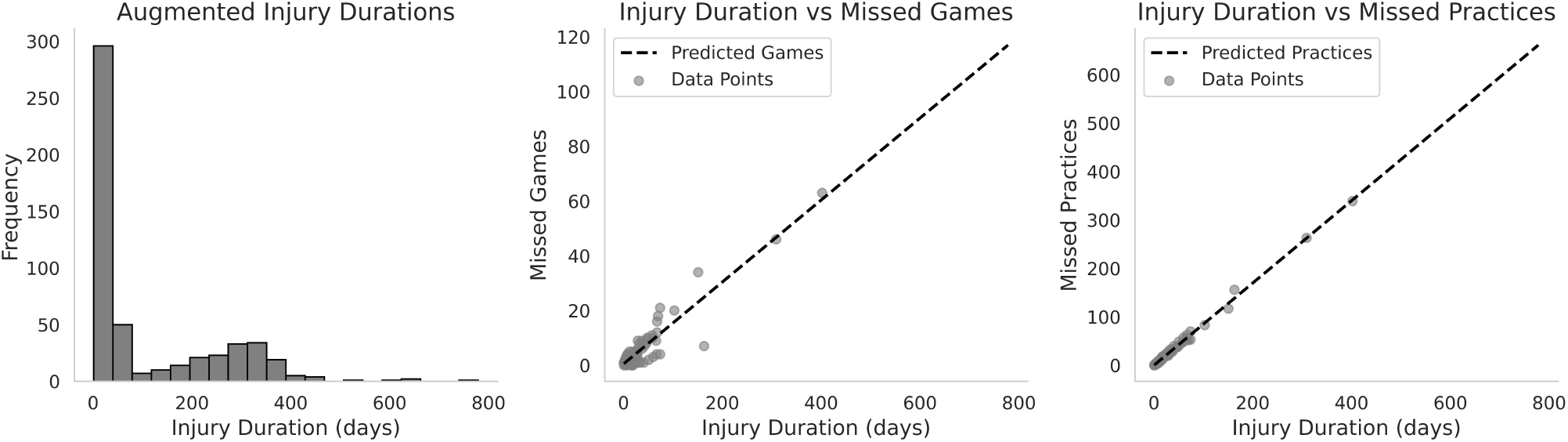
Augmented Injury Duration Data. The figure shows the Distribution of the augmented injury data as well as an exemplarily fit of the number of missed games as well as missed practices dependent on the injury duration.

**Figure S9:**
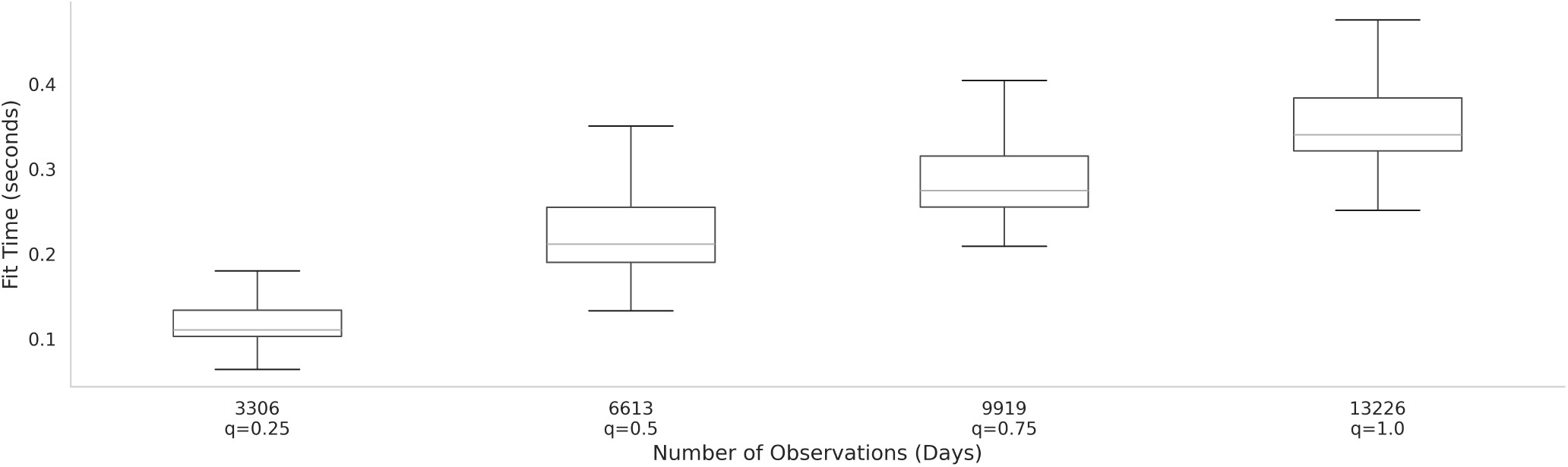
Fitting Times in Seconds of the XGBSE Model. The plot shows the fitting times of the XGBSE model on 25%, 50%, 75%, and 100% of the data. Fitting was repeated 400 times. If *q <* 1, the data was sampled randomly with stratification for each of the 400 iterations.

## Supplementary Tables

**Table S1:**
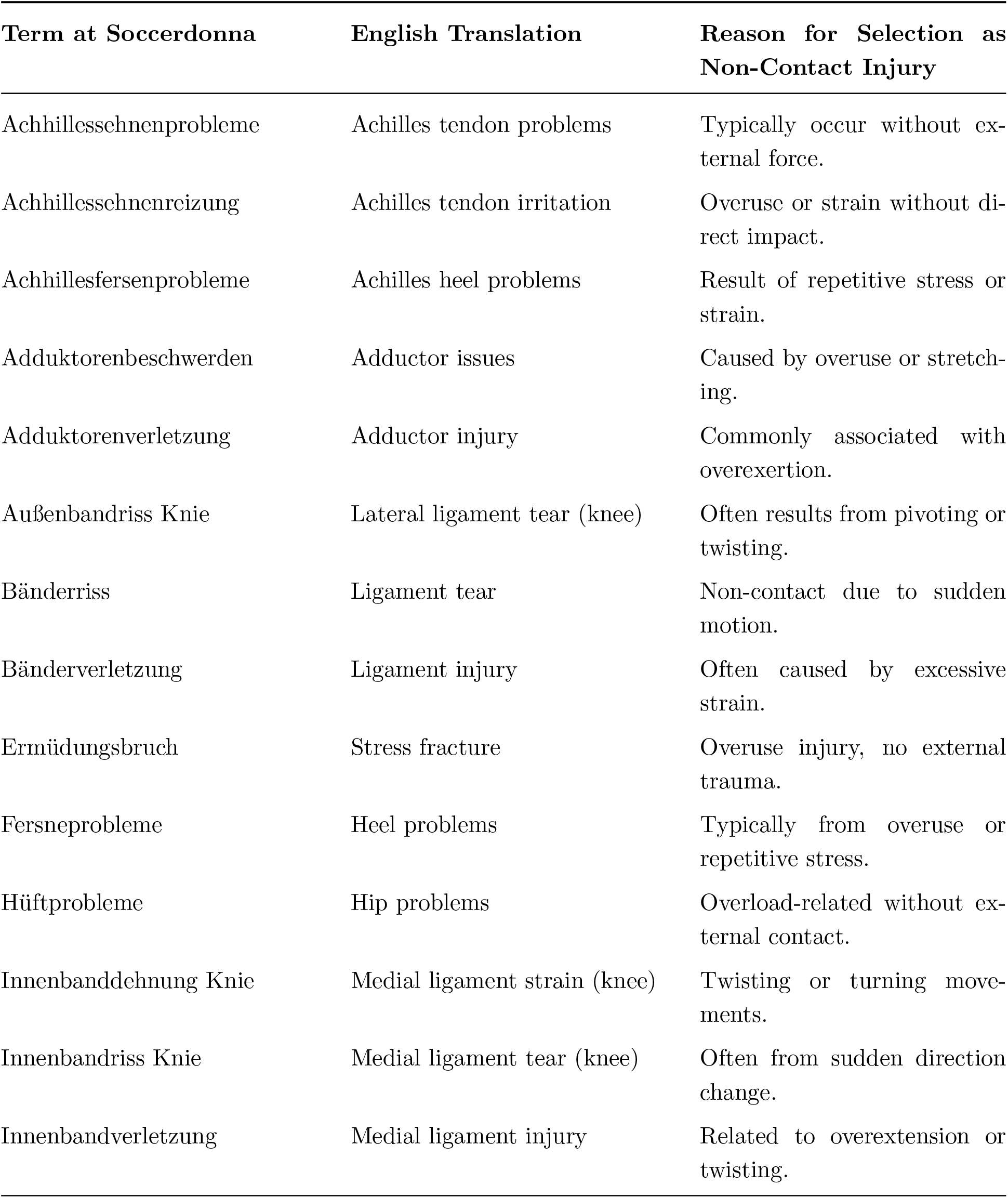
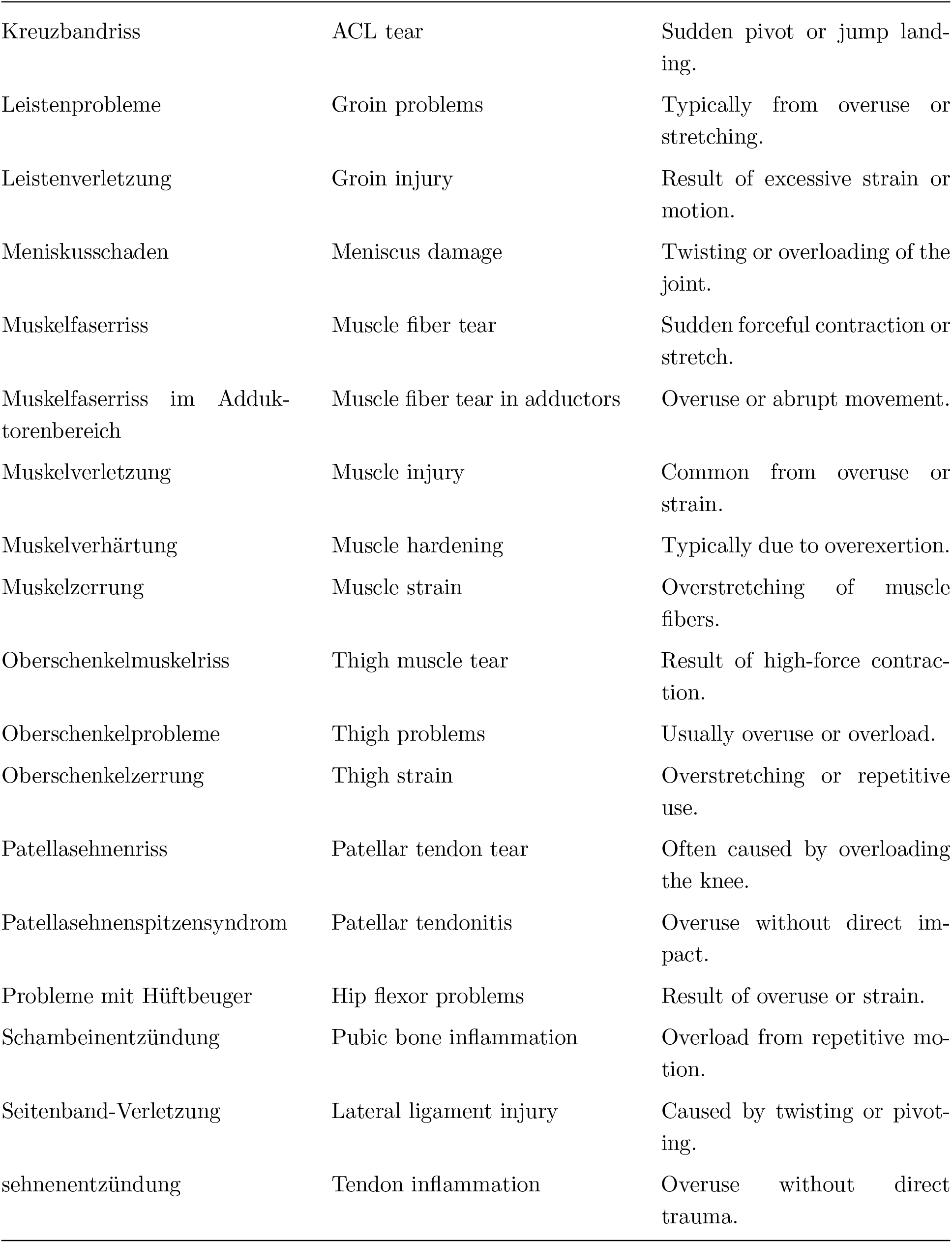
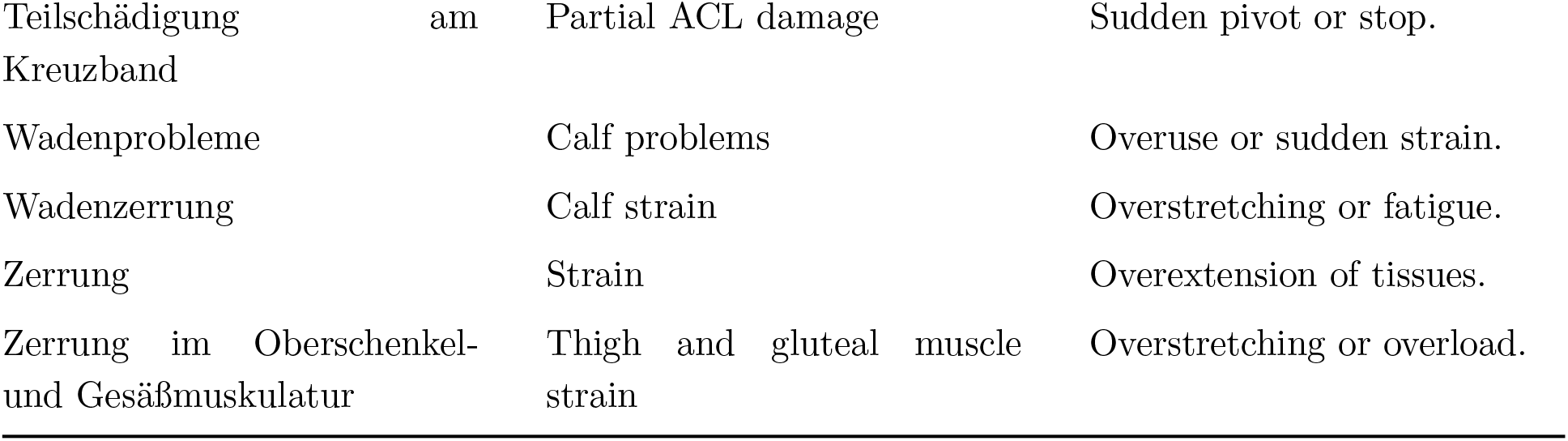
Scraped Non-Contact Injury Types. The table represents all injuries we used from Soccerdonna^39^ in order to augment the injury duration data set as outlined within the Method section.

**Table S2:** Rules for data pre-processing. The table summarizes how missing training information was handled during feature engineering, as described in the Method section.

|  | Handling |
| --- | --- |
| **Physical Activity** |  |
| A day at which the total running distance covered is greater than zero at a specific day | Reported value |
| **Rest day** |  |
| A day at which the total running distance is not reported, with a match on the previous day | Zero training load |
| A day at which the total running distance is not reported, flanked by non-zero values on both the previous and next day | Zero training load |
| 2 (3) consecutive days for which the total running distance is not reported and is preceded and succeeded by a reported day | Zero training load |
| The sequence of days for which the total running distance is not reported including the 24th of December | Zero training load |
| The sequence of days for which the total running distance is not reported including the 1st of January | Zero training load |
| The sequence of days for which the total running distance is not reported including the 1st of July 2020 and 2022 | Zero training load |
| **National team** |  |
| The sequence of days for which the total running distance is not reported but one of the days is associated with a national match | Imputed but only used as information for previous exposure |
| **Injury recovery** |  |
| The sequence of days from the day after an injury to 3 weeks after the first day of recorded physical activity after the injury | Dropped |
| **Unknown activity** |  |
| Days for which none of the other rules apply to. Includes COVID19 pandemic period. | Dropped |

**Table S3:** Engineered Features Description. The table describes the engineered features that were used for model training, as described in the Method section.

| Feature Name | Description | Calculation Method |
| --- | --- | --- |
| **Categorized Training Load** |  |  |
| Distance Covered per Minute | Distance per Minute covered in meters during training or match play. | Directly measured using player tracking data. |
| High Metabolic Load Distance (HMLD) per Minute | Distance per Minute covered when metabolic power exceeds 25.5 W/kg, typically occurring at speeds above 5.5 m/s. | Directly measured using player tracking data. |
| Number of Accelerations per Minute | Number of acceleration events per Minute. | Directly measured using player tracking data. |
| Number of Decelerations per Minute | Number of deceleration events per Minute. | Directly measured using player tracking data. |
| Distance Covered during Acceleration per Minute | Distance covered per Minute during acceleration events. | Directly measured using player tracking data. |
| Distance Covered during Deceleration per Minute | Distance covered per Minute during deceleration events. | Directly measured using player tracking data. |
| High-Speed Running Distance per Minute (Relative HSR) | Distance covered per Minute while running at speeds above 18 km/h. | Directly measured using player tracking data. |
| **Accumulated Training Load** |  |  |

**Table S3 – continued from previous page**
| Feature Name | Description | Calculation Method |
| --- | --- | --- |
| EWMA | Exponentially Weighted Moving Average (EWMA) of past training loads. Provides a smoothed estimate with greater weight on recent loads. | $\alpha e_{i,t} + (1 - \alpha)EWMA_{i,t-1}^j$<br>with $\alpha = 2/11$ as in<sup>6</sup> |
| ACWR | Acute Chronic Workload Ratio (ACWR) of the respective training loads. Assesses the balance between acute and chronic load, indicating potential injury risk. | $\frac{\text{Acute Load (last 6 days)}}{\text{Chronic Load (last 28 days)}}$ |
| MWR | Monotonic Workload Ratio (MWR) of the respective training loads. Evaluates training load stability by considering the mean and standard deviation over the last 7 days. | $\frac{\text{Mean of Last 7 Days}}{\text{Standard Deviation of Last 7 Days}}$ |

**Table S4:** Hyperparameter Search Spaces. This table summarizes the hyperparameter search spaces for each classification model that was used as comparison against the survival-based models. They were applied with Bayesian hyperparameter optimization with 60 iterations for each model. Integer[a,b] indicates all integers values from a to b. Real[a,b] indicates all real values from a to b. Categorical[a,b,c] indicates the values a,b, and c. The option log-uniform indicates that the respective values are drawn uniformly from the logarithm of the bounds.

| Model Name | Hyperparameter | Range/Type |
| --- | --- | --- |
| **k-Nearest Neighbors (KNN)** |  |  |
| Number of Neighbors ($n_{neighbors}$) | Number of nearest neighbors to consider for classification or regression. | Integer [3, 20] |
| Weights | Weight function used in prediction. | Categorical ['uniform', 'distance'] |
| Distance Metric ($p$) | Distance metric for the tree (Manhattan or Euclidean). | Categorical [1, 2] |
| **LightGBM** |  |  |
| Number of Estimators ($n_{estimators}$) | Number of boosting iterations. | Integer [20, 300] |

Table S4 – continued from previous page
| Model Name | Hyperparameter | Range/Type |
| --- | --- | --- |
| Maximum Depth<br>(*max_depth*) | Maximum depth of the tree. | Integer [3, 8] |
| Learning Rate<br>(*learning_rate*) | Shrinkage parameter for boosting. | Real [0.1, 0.5] |
| Subsample (*subsample*) | Fraction of samples used for fitting. | Real [0.5, 1.0] |
| Column Subsampling<br>(*colsample_bytree*) | Fraction of columns sampled for each tree. | Real [0.5, 1.0] |
| **LightGBM with Monotone Activity Duration** |  |  |
| Number of Estimators<br>(*n_estimators*) | Number of boosting iterations. | Integer [20, 200] |
| Maximum Depth<br>(*max_depth*) | Maximum depth of the tree. | Integer [3, 8] |
| Learning Rate<br>(*learning_rate*) | Shrinkage parameter for boosting. | Real [0.1, 0.5] |
| Subsample (*subsample*) | Fraction of samples used for fitting. | Real [0.5, 1.0] |
| Column Subsampling<br>(*colsample_bytree*) | Fraction of columns sampled for each tree. | Real [0.5, 1.0] |
| **Logistic Regression (LR)** |  |  |
| Regularization Parameter<br>(*C*) | Inverse of regularization strength. | Real [1e-4, 1e2, log-uniform] |
| **LightGBM with Tree Embedding** |  |  |
| Number of Estimators<br>(*n_estimators*) | Number of boosting iterations (LightGBM). | Integer [20, 300] |
| Maximum Depth<br>(*max_depth*) | Maximum depth of the tree (LightGBM). | Integer [3, 8] |
| Learning Rate<br>(*learning_rate*) | Shrinkage parameter for boosting (LightGBM). | Real [0.1, 0.5] |
| Subsample (*subsample*) | Fraction of samples used for fitting (LightGBM). | Real [0.5, 1.0] |
| Column Subsampling<br>(*colsample_bytree*) | Fraction of columns sampled for each tree (LightGBM). | Real [0.5, 1.0] |

**Table S4 – continued from previous page**
| **Model Name** | **Hyperparameter** | **Range/Type** |
| --- | --- | --- |
| Regularization Parameter<br>($C$) | Inverse of regularization strength (Logistic Regression). | Real [1e-4, 1e2, log-uniform] |

## References

[1] Martin Hägglund, Markus Waldén, Henrik Magnusson, Karolina Kristenson, Håkan Bengtsson, and Jan Ekstrand. Injuries affect team performance negatively in professional football: an 11-year follow-up of the uefa champions league injury study. British journal of sports medicine, 47(12):738–742, 2013.

[2] Eyal Eliakim, Elia Morgulev, Ronnie Lidor, and Yoav Meckel. Estimation of injury costs: financial damage of english premier league teams’ underachievement due to injuries. BMJ Open Sport & Exercise Medicine, 6(1):e000675, 2020.

[3] Olivia A Hurley. Impact of player injuries on teams’ mental states, and subsequent performances, at the rugby world cup 2015. Frontiers in psychology, 7:807, 2016.

[4] SR Filbay, AG Culvenor, IN Ackerman, TG Russell, and KM Crossley. Quality of life in anterior cruciate ligament-deficient individuals: a systematic review and meta-analysis. British Journal of Sports Medicine, 49(16):1033–1041, 2015.

[5] Howden. Howden injury index: Sports injury trends and insights, 2023. Available at: https://www.howdengroup.com.

[6] Alessio Rossi, Luca Pappalardo, Paolo Cintia, F Marcello Iaia, Javier Fernández, and Daniel Medina. Effective injury forecasting in soccer with gps training data and machine learning. PLOS One, 13(7):e0201264, 2018.

[7] Alejandro López-Valenciano, Francisco Ayala, José Miguel Puerta, Mark De Ste Croix, Francisco Vera-García, Sergio Hernández-Sánchez, Iñaki Ruiz-Pérez, and Gregory Myer. A preventive model for muscle injuries: a novel approach based on learning algorithms. Medicine and science in sports and exercise, 50(5):915, 2018.

[8] Francisco Ayala, Alejandro López-Valenciano, Jose Antonio Gámez Martín, Mark De Ste Croix, Francisco J Vera-Garcia, Maria del Pilar Garcia-Vaquero, Iñaki Ruiz-Pérez, and Gregory D Myer. A preventive model for hamstring injuries in professional soccer: Learning algorithms. International journal of sports medicine, 40(05):344–353, 2019.

[9] Nikki Rommers, Roland Rössler, Evert Verhagen, Florian Vandecasteele, Steven Verstockt, Roel Vaeyens, Matthieu Lenoir, Eva D’Hondt, and Erik Witvrouw. A machine learning approach to assess injury risk in elite youth football players. Medicine and science in sports and exercise, 52(8):1745–1751, 2020.

[10] Garrett S Bullock, Joseph Mylott, Tom Hughes, Kristen F Nicholson, Richard D Riley, and Gary S Collins. Just how confident can we be in predicting sports injuries? a systematic review of the methodological conduct and performance of existing musculoskeletal injury prediction models in sport. Sports medicine, 52(10):2469–2482, 2022.

[11] Christopher Leckey, Nicol van Dyk, Cailbhe Doherty, Aonghus Lawlor, and Eamonn Delahunt. Machine learning approaches to injury risk prediction in sport: a scoping review with evidence synthesis. British Journal of Sports Medicine, 2024.

[12] Jon L Oliver, Francisco Ayala, Mark BA De Ste Croix, Rhodri S Lloyd, Greg D Myer, and Paul J Read. Using machine learning to improve our understanding of injury risk and prediction in elite male youth football players. Journal of science and medicine in sport, 23(11):1044–1048, 2020.

[13] Emmanuel Vallance, Nicolas Sutton-Charani, Abdelhak Imoussaten, Jacky Montmain, and Stéphane Perrey. Combining internal-and external-training-loads to predict non-contact injuries in soccer. Applied Sciences, 10(15):5261, 2020.

[14] Ewout W Steyerberg, Andrew J Vickers, Nancy R Cook, Thomas Gerds, Mithat Gonen, Nancy Obuchowski, Michael J Pencina, and Michael W Kattan. Assessing the performance of prediction models: a framework for traditional and novel measures. Epidemiology, 21(1):128–138, 2010.

[15] Chuan Guo, Geoff Pleiss, Yu Sun, and Kilian Q Weinberger. On calibration of modern neural networks. In International conference on machine learning, pages 1321–1330. PMLR, 2017.

[16] Davi Vieira, Gabriel Gimenez, Guilherme Marmerola, and Vitor Estima. Xgboost survival embeddings: improving statistical properties of xgboost survival analysis implementation, 2021. URL https://loft-br.github.io/xgboost-survival-embeddings/.

[17] Avinash Barnwal, Hyunsu Cho, and Toby Hocking. Survival regression with accelerated failure time model in xgboost. Journal of Computational and Graphical Statistics, 31 (4):1292–1302, 2022.

[18] Noah Simon, Jerome H Friedman, Trevor Hastie, and Rob Tibshirani. Regularization paths for cox’s proportional hazards model via coordinate descent. Journal of statistical software, 39:1–13, 2011.

[19] Meelis Kull, Telmo Silva Filho, and Peter Flach. Beta calibration: a well-founded and easily implemented improvement on logistic calibration for binary classifiers. In Artificial intelligence and statistics, pages 623–631. PMLR, 2017.

[20] Monika E Maros, David Capper, David T W Jones, Volker Hovestadt, Andreas von Deimling, Stefan M Pfister, Martin Sill, and Felix Sahm. Machine-learning workflows for the classification of dna methylation array data. Nature Protocols, 15(2):479–512, 2019. doi: 10.1038/s41596-019-0251-6.

[21] Haoyue Huang, Jiawei Zheng, Hao Zhang, Bing Wang, Mingxi Zhou, Jiacheng Zhang, and Ying Wang. Machine learning-based tissue of origin classification for cancers of unknown primary. Nature Communications, 13(1):4327, 2022. doi: 10.1038/s41467-022-31666-w.

[22] Gil Rodas, Lourdes Osaba, David Arteta, Ricard Pruna, Dolors Fernández, and Alejandro Lucia. Genomic prediction of tendinopathy risk in elite team sports. International Journal of Sports Physiology and Performance, 15(4):489–495, 2019.

[23] Juan Ramon González, Alejandro Cáceres, Eva Ferrer, Laura Balagué-Dobón, Xavier Escriba-Montagut, David Sarrat, Guillermo Quintás, and Rodas Rodas. Predicting injuries in elite female football playeߪ International Journal of Sports Physiology and Performance, 19:661–669, 2024.

[24] Ricard Pruna, Thor Einar Andersen, Ben Clarsen, and Alan McCall. Muscle injury guide: Prevention of and return to play from muscle injuries. Technical report, Barca Innovation Hub, 2018.

[25] Jill Cook, Gil Rodas, Alan McCall, Ricard Pruna, Rochelle Kennedy, and Lluís Til. Tendon Injuries in Football Players: FC Barcelona 2021 Tendon Guide. FC Barcelona: Barça Innovation Hub, Barcelona, Spain, 1st edition, 2021.

[26] Guolin Ke, Qi Meng, Thomas Finley, Taifeng Wang, Wei Chen, Weidong Ma, Qiwei Ye, and Tie-Yan Liu. Lightgbm: A highly efficient gradient boosting decision tree. Advances in Neural Information Processing Systems, 30, 2017.

[27] Jorma Laaksonen and Erkki Oja. Classification with learning k-nearest neighbors. In Proceedings of international conference on neural networks (ICNN’96), volume 3, pages 1480–1483. IEEE, 1996.

[28] David W Hosmer Jr, Stanley Lemeshow, and Rodney X Sturdivant. Applied logistic regression. John Wiley & Sons, 2013.

[29] Ahmed Naglah, Fahmi Khalifa, Ali Mahmoud, Mohammad Ghazal, Paul Jones, Teena Murray, Adel S Elmaghraby, and Ayman El-Baz. Athlete-customized injury prediction using training load statistical records and machine learning. In 2018 IEEE international symposium on signal processing and information technology (ISSPIT), pages 459–464. IEEE, 2018.

[30] Greg Ridgeway. The state of boosting. Computing science and statistics, pages 172–181, 1999.

[31] Toshihito Takahashi, Kazunori Nozaki, Tomoya Gonda, Tomoaki Mameno, and Kazunori Ikebe. Deep learning-based detection of dental prostheses and restorations. Scientific Reports, 11(1):1960, 2021.

[32] Todd Hollon, Cheng Jiang, Asadur Chowdury, Mustafa Nasir-Moin, Akhil Kondepudi, Alexander Aabedi, Arjun Adapa, Wajd Al-Holou, Jason Heth, Oren Sagher, et al. Artificial-intelligence-based molecular classification of diffuse gliomas using rapid, label-free optical imaging. Nature medicine, 29(4):828–832, 2023.

[33] Cox R David et al. Regression models and life tables (with discussion). Journal of the Royal Statistical Society, 34(2):187–220, 1972.

[34] Sebastian Pölsterl. scikit-survival: A library for time-to-event analysis built on top of scikit-learn. Journal of Machine Learning Research, 21(212):1–6, 2020.

[35] Tianqi Chen and Carlos Guestrin. XGBoost: A scalable tree boosting system. In Proceedings of the 22nd ACM SIGKDD International Conference on Knowledge Discovery and Data Mining, KDD ‘16, pages 785–794, New York, NY, USA, 2016. ACM. ISBN 978-1-4503-4232-2.

[36] Xinran He, Junfeng Pan, Ou Jin, Tianbing Xu, Bo Liu, Tao Xu, Yanxin Shi, Antoine Atallah, Ralf Herbrich, Stuart Bowers, et al. Practical lessons from predicting clicks on ads at facebook. In Proceedings of the eighth international workshop on data mining for online advertising, pages 1–9, 2014.

[37] F. Pedregosa, G. Varoquaux, A. Gramfort, V. Michel, B. Thirion, O. Grisel, M. Blondel, P. Prettenhofer, R. Weiss, V. Dubourg, J. Vanderplas, A. Passos, D. Cournapeau, M. Brucher, M. Perrot, and E. Duchesnay. Scikit-learn: Machine learning in Python. Journal of Machine Learning Research, 12:2825–2830, 2011.

[38] Tim Head, Manoj Kumar, Holger Nahrstaedt, Gilles Louppe, and Iaroslav Shcherbatyi. scikit-optimize/scikit-optimize, October 2021.

[39] Soccerdonna. Soccerdonna - women’s soccer database. https://www.soccerdonna.de.

[40] Marc Guitart, Martí Casals, David Casamichana, Jordi Cortés, Francesc Xavier Valle, Alan McCall, Francesc Cos, and Gil Rodas. Use of gps to measure external load and estimate the incidence of muscle injuries in men’s football: A novel descriptive study. PLOS One, 17(2):e0263494, 2022.

[41] Donald R. Jones, Matthias Schonlau, and William J. Welch. Efficient global optimization of expensive black-box functions. Journal of Global Optimization, 13(4):455–492, 1998. doi: 10.1023/A:1008306431147.

[42] Jasper Snoek, Hugo Larochelle, and Ryan P Adams. Practical bayesian optimization of machine learning algorithms. Advances in neural information processing systems, 25, 2012.

[43] World Medical Association. Wma declaration of helsinki – ethical principles for medical research involving human subjects, 2013. Retrieved from https://www.wma.net/policies-post/wma-declaration-of-helsinki-ethicalprinciples-for-medical-research-involving-human-subjects/.

